# Vaccination under the Pandemic and Political Support

**DOI:** 10.1101/2022.02.13.22270703

**Authors:** Masaki Takahashi, Reo Takaku, Toyo Ashida, Yoko Ibuka

## Abstract

This paper examines the impact of COVID-19 vaccination on political support. We exploit Japan’s age-based COVID-19 vaccine roll-out, prioritizing people aged 65 years and older. A regression discontinuity design based on a large-scale online survey reveals that COVID-19 vaccination increases favorable opinions of the vaccination progress in municipalities and infection control measures of municipal governments. By contrast, there is no significant effect on support for the central government. We also discovered that people with chronic diseases and low socioeconomic status are more affected. These results show that reciprocal motives and political information play an important role in evaluating public policies.

## 1 Introduction

The interaction between government policies and political support has been a central issue in both political science (Dahlberg and Johansson, 2002; Rudolph and Evans, 2005; De La O, 2013; Marshall, 2019) and public economics (Manacorda et al., 2011; Linos, 2013; Labonne, 2013). Under a public health crisis like the COVID-19 pandemic, the government’s infection control measures directly affect people’s lives and livelihoods, and these policies attract attention from the public. News headlines worldwide have been dominated by how many people are infected and killed by COVID-19, how the government responds to it, and what it will take to overcome the pandemic. As a result, many studies have examined the intended and unintended consequences of government policies against the pandemic, such as social distancing measures and economic stimulus packages. However, despite the high profile of the government policies against the pandemic, their political consequences have remained largely unknown.^1^

This paper explores how COVID-19 vaccination affects people’s political support for the government. Vaccination is one of the most effective interventions among many measures to control the pandemic. The timing of vaccine approval, the availability of vaccines, and the speed of vaccine dissemination are all strongly linked to government policies and thus politics (Bokemper et al., 2021). For example, 78% of Americans believe that the COVID-19 vaccine approval process was driven more by politics than science (Meyer, 2020). Furthermore, so-called “vaccine nationalism” has prevailed among developed countries, as to how well the government procured COVID-19 vaccines for its own country was regarded as an indication of its political competence (Fidler, 2020; Katz et al., 2021). While COVID-19 vaccination was accelerated due to political motivation in developed countries, it is unclear whether vaccination has led to political support for the government.

It is necessary to find the exogenous variations of vaccination status among the population to estimate the causal impact of COVID-19 vaccination on political support. However, it is difficult to find such a situation in practice because self-selection largely influences vaccination uptake. We exploit an age-based COVID-19 vaccine roll-out in Japan to overcome this difficulty. Older people are at higher risk of hospitalization and death when infected with COVID-19 (CDC, 2022). Therefore, like many other countries, the Japanese government prioritized older people to receive the vaccine. Specifically, the elderly born before April 1, 1957, who have reached or will reach 65 years of age during the fiscal year 2021, could be vaccinated before younger people. This age-based prioritization has resulted in exogenous variation in vaccination rates around the age 65 threshold during the vaccine dissemination phase. Our study exploits the sharp variation in vaccination rates at the age cut-off to estimate the political impact of COVID-19 vaccination.

We conducted an online survey to obtain information on people’s vaccination status and political opinions. Our survey was conducted in August 2021, when the gap in vaccination rates between those aged 65 and older and those under 65 was the highest. The survey responses were collected from 30,892 elderly individuals who were around the cut-off age of 65 years. In addition to vaccination status and political opinions, we also collected various information on respondents’ demographics, health behaviors, and health outcomes. This information enables us to examine which demographics are associated with the political impact of vaccination and how vaccination affects people’s health behavior and health outcomes.

Using the survey data, we adopt a fuzzy regression discontinuity (RD) design to estimate the impact of COVID-19 vaccination on political support for the municipal and central governments. The first stage estimates indicate that at the age 65 threshold, the vaccination rate jumps by 29.3 percentage points. Our fuzzy RD estimation reports the following main results based on the significant first-stage impact. First, COVID-19 vaccination significantly increases favorable opinions of vaccination progress in municipalities and municipal government infection control measures. However, there is no significant effect on support for the central government. The difference in the impact between municipal and central government is likely to reflect the institutional setting in Japan, where municipal governments play a primary role in establishing the vaccine delivery system for older people.

Second, when respondents are classified by health condition based on the presence or absence of chronic diseases, those with chronic diseases are more likely to increase political support due to vaccination. It is widely known that people with pre-existing health conditions are at greater risk of hospitalization and death from infection with COVID-19. Therefore, those with chronic diseases might be more appreciative of vaccination than those without and take their vaccination status as crucial political information. We also find that people with a high level of interpersonal trust increase their support for the government more than those with low trust levels. Interpersonal trust is closely associated with reciprocity (Ostrom and Walker, 2003; Altmann et al., 2008)^2^. These results suggest that people increase their political support for the government through reciprocal motivation toward the government.

Third, people with lower socioeconomic status (SES) are more likely to increase political support through vaccination. Specifically, the political impact of vaccination is more pronounced among those with low income and those without a college degree. One interpretation of these results is that people with lower SES have limited knowledge about the government policies (Angelucci and Prat, 2021). Therefore, these people may be more likely to update their political knowledge about the government policies when they benefit from them. These results are also consistent with theoretical predictions about voter behavior when they have limited information about government abilities (Rogoff, 1990; Manacorda et al., 2011).

This paper contributes to the literature that analyzes the interaction between government policy and political support using one of the most important public health policies during the COVID-19 pandemic: vaccination. We take advantage of the fact that COVID-19 vaccination is a universal public policy that targets the general population. By analyzing the political responses of people with different health statuses, personal values, and SES, we examine the mechanisms by which people increase their political support for governments that provide public services. Considering that most previous studies have focused on people with specific SES and had limited statistical power to explore the mechanisms, examining multiple political support mechanisms is an important contribution of this study.

Previous studies have focused mainly on the political impact of public policies targeting people from specific socioeconomic backgrounds, especially the poor population. Manacorda et al. (2011) and De La O (2013) respectively examine the impact of cash transfer to the poor people on their political support for the government in Uruguay and Mexico.^3^ These studies find that government transfers to the disadvantaged people significantly increase their political support for the government. Manacorda et al. (2011) argues that the increased political support among poor people is mainly attributed to the insufficient information about the competence of the incumbent, as indicated by a model of rational but poorly informed voters (Rogoff, 1990).^4^ Furthermore, other studies indicate the importance of pure psychological motivations among voters, including the reciprocal relationship between voters and politicians (Hahn, 2009; Finan and Schechter, 2012), rather than rational decision making among voters (Rogoff, 1990). Our estimation using individual attributes indicates the importance of both political information and reciprocal motives in people’s political support.

This paper is organized as follows. Section 2 presents the age-based priority policy for COVID-19 vaccination in Japan. Section 3 discusses mechanisms of why COVID-19 vaccination can affect people’s political opinions. Section 4 describes our survey data and presents descriptive statistics. Section 5 explains empirical strategies applying RD design. Section 6 reports estimation results for the political impact of COVID-19 vaccination. Section 7 discusses other potential effects of COVID-19 vaccination and presents estimation results for robustness checks. Section 8 concludes this paper.

## 2 Background

### 2.1 Age-based Priority Rule and Vaccination Trends

This section describes the COVID-19 vaccine delivery system in Japan. As in many countries, vaccination was considered to be particularly effective in controlling COVID-19 infection in Japan. Against the backdrop of the public expectations for vaccines, the Japanese government made the spread of the vaccine a top priority in the fight against COVID-19. However, the spread of vaccines in Japan lagged behind other developed countries because of the slow progress of regulatory approval (Matsui et al., 2021; Kosaka et al., 2021). After the vaccines were approved,^5^ the Japanese government rushed to purchase the vaccine from pharmaceutical companies, and vaccination for the general public began in April 2021.

A significant feature of the COVID-19 vaccination policy in Japan is the age-based priority, which is called the “Older-people-first Policy” (Matsui et al., 2021).^6^ The Japanese government decided to give priority to older people to receive the vaccine because they are known to be at higher risk of severe disease and death when infected with COVID-19. Specifically, the Japanese government has allowed older people born before April 1, 1957, who have reached or will reach the age of 65 years during the fiscal year 2021, to be vaccinated before younger people.^7^ Following this priority, each municipal government allowed older people to receive the vaccine before the younger ones. Municipalities that have finished vaccinating older people will proceed to vaccinate younger ones.

Although the diffusion of vaccines in Japan lagged behind other developed countries, the central government began mass vaccination programs in earnest. Prime Minister Yoshihide Suga promised the public on May 13 to vaccinate 36 million elderly citizens aged 65 years and older by the end of July 2021.^8^. Starting in early June 2021, vaccination rates began to rise rapidly. Figure 1 plots the vaccination rate for the second dose among the population aged 65 years and older and those under 65 years, based on publicly available government official data.

**Figure 1:**
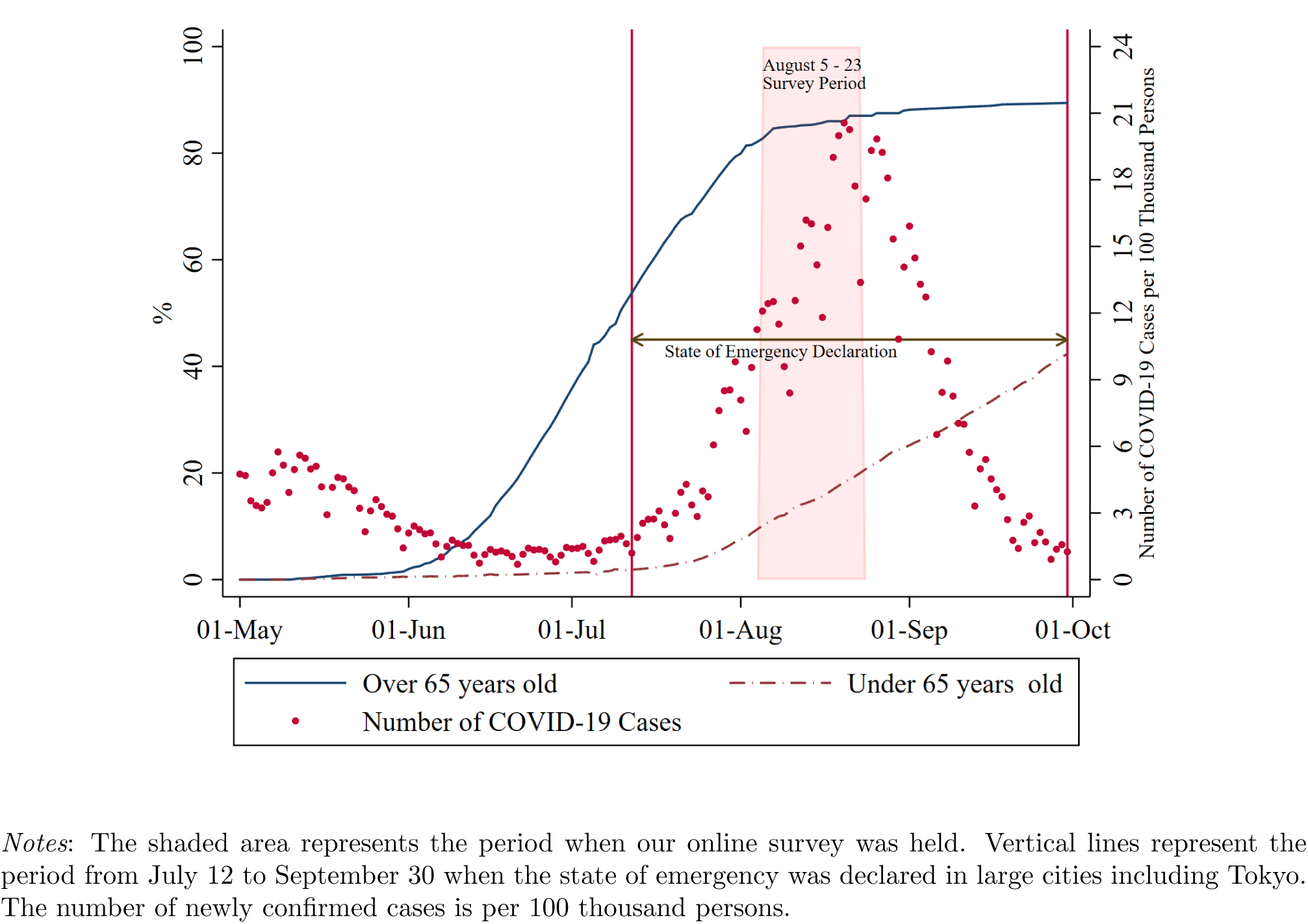
Trends of Second Dose Vaccination and Number of Infected People

Among people aged 65 years and older, vaccination rates have increased from 0.1 percent on June 1 to 79 percent on July 31. Among those under 65 years, the diffusion of vaccines lagged for the elderly population because of the priority rule based on the threshold of age 65.^9^

The difference between vaccination rates of the second dose among people aged 65 years and older and those under 65 years was maximized in early August. On August 6, the next day when our original survey began, the vaccination rate of the second dose among people aged 65 years and older and under 65 years was 83% and 11%, respectively.

Figure 1 also shows the number of infected people per 100 thousand (red dots). While vaccination of the elderly had increased since early June, COVID-19 infection increased in August because of the spread of the Delta variant. To control the infection, the state of emergency was declared in Tokyo on July 12 and was extended to 21 prefectures in late August, the peak of the outbreak.^10^ Eventually, the state of emergency was lifted on September 30, following a rapid decrease in the number of infected people.

### 2.2 Role of Municipalities in Vaccination Progress

While the central government is responsible for securing and distributing the vaccine to municipalities, each municipal government administers the vaccination.^11^ First, each municipal government is responsible for preparing vaccination sites and providing vaccines based on the priority. The municipal governments worked with local hospitals and clinics to prepare medical facilities and mass vaccination sites where vaccinations could be given. For residents to make an appointment for vaccination, they need to receive a coupon sent by the municipal government. Second, each municipality is responsible for securing human resources for vaccination in cooperation with the local medical association and needs to provide a high daily allowance for doctors and nurses. In Japan, only doctors and nurses are legally allowed to inject the COVID-19 vaccine, so the lack of human resources has been a significant obstacle to the distribution of the vaccine. Many municipalities had difficulty in securing enough qualified human resources for COVID-19 vaccination because doctors and nurses must also provide their regular medical care (Kosaka et al., 2021).

Reflecting on the crucial role of municipalities in vaccination progress, there was a significant regional variation in vaccination rates. Appendix Figure A1 show the prefecture-level map on the difference of the vaccination rate from the mean value for each age group. In these figures, we confirm modest regional disparities in the vaccination rate of the second dose. For example, there appears to be a significant disparity in vaccination rates in the middle of mainland Japan.

## 3 COVID-19 Vaccination and Political Support

### 3.1 Reciprocal Motive

This section explains why COVID-19 vaccination can influence people’s political opinions. Previous studies have shown that reciprocal relationships between voters and politicians play a crucial role in the political process (Finan and Schechter, 2012; Hahn, 2009).^12^ If the government policy is favorable to some voters, the beneficiaries may feel grateful and may want to reward the government. In the case of COVID-19 vaccination, people may increase their support for the government in gratitude for being given priority vaccination or taking the vaccine. Since it is generally difficult to directly measure the intrinsic reciprocity of individuals, we explore its importance in political support from the following two perspectives.

#### Chronic Disease

An important feature of COVID-19 is that people with certain diseases are more likely to get severely ill from the infection. Therefore, vaccination has significant benefits, especially for those with underlying medical conditions. Given that the benefits of vaccines vary by health status, vaccination may be more likely to increase political support among people with chronic diseases because of reciprocal motives toward the government.

#### Interpersonal Trust

Considering reciprocal motives, a person with a personality that values reciprocity might be more likely to increase political support for the government due to vaccination. Here, we pay attention to people’s interpersonal trust. Our hypothesis is that people with a high level of interpersonal trust also have high reciprocity toward the government, and that such people are more likely to increase their support for the government by receiving public services. Reciprocity is a crucial aspect of interpersonal trust (Ostrom and Walker, 2003; Fukuyama, 1996).^13^ As Ostrom and Walker (2003) pointed out that “the levels of trust and reciprocity and reputations for being trustworthy are positively reinforcing”, it is plausible to expect a positive relationship between interpersonal trust and reciprocity. Several empirical studies revealed a close relationship between them. For example, Altmann et al. (2008) conducted a laboratory experiment and find a strong positive correlation between a person’s reciprocity and her trusting behavior.^14^

### 3.2 Political Information and Socio-Economic Status

In addition to reciprocity, people’s political information could also play an important role in their political support. Modern political economy models have formulated the relationship between government policy and political support as voters’ learning about the government’s capabilities (Rogoff, 1990; Manacorda et al., 2011). Because of various information constraints, voters cannot understand the actual consequences of government policies. Under this asymmetric information, voters use the related outcomes as signals to infer them.

To put it in the context of COVID-19 vaccines, people may infer the government’s ability on vaccine policies and other infection control measures based on whether they were able to get vaccinated. The above theoretical model implies that whether vaccination leads to support for the government policies depends on how much people know about policies in advance. For example, if people knew the actual vaccination progress in their municipalities and nationwide, vaccination would not influence their political support. Meanwhile, people may not know vaccination progress in their municipalities and across the country. They may also overestimate their vaccination status and use this information as a signal to infer overall vaccination progress. In this case, getting vaccinated can lead to support for the government.

The importance of political information also suggests that the political impact of vaccination depends on personal attributes such as socioeconomic status (SES). For example, less educated people may not have accurate information about the prevalence of vaccines and may be more likely to change their support for the government based on their vaccination status. Angelucci and Prat (2021) investigate information inequality about political news by SES groups. They find that people with high SES have more accurate information than those with low SES and that socioeconomic factors matter more than partisanship in determining information levels. Therefore, we also analyze which socioeconomic factors drive the political impact of COVID-19 vaccination.

## 4 Data

### 4.1 Online Survey

To gather the necessary information for our analysis, we hired an internet-survey company called Cross Marketing Inc., one of the largest survey companies in Japan. They employed random sampling from about 4,790,000 people across the nation who had preregistered as potential survey participants. Our survey was conducted from August 5 to 23, 2021, around the time when the difference in vaccination rates between people aged 65 years and older and those under 65 years was greatest, as shown in Figure 1. We targeted people born between April 1954 and March 1960, in other words, those between the ages of 62 and 67 years at the end of the fiscal year 2021 (March 2022). The survey was sent to 54,356 married survey registrants, and we gathered 30,892 responses. Our survey asked a wide range of questions related to vaccination, political opinions, health awareness and behaviors, and SES. In the following, we explain the main variables used in the analysis.

### 4.2 Variable of Interest

#### Vaccination status

The COVID-19 vaccine is supposed to be given twice, three to four weeks apart. We asked the survey participants how many times and when they had been vaccinated. If they had not been vaccinated, we asked whether they were planning to be vaccinated. In estimation, we use whether a respondent has completed two doses of COVID-19 vaccination as the variable of interest (vaccination status) because it is widely believed that completing two doses is necessary to develop an antibody.

### 4.3 Dependent Variables

#### Political opinions

The main outcome variable in our analysis is people’s political opinions related to the pandemic. As discussed in Section 2, the central government and municipal government played different roles in the supply of vaccines in Japan. Therefore, we asked respondents about their political opinions of both the central government and the municipal government.

We measured respondents’ support for the government by asking their opinions about government policies against the pandemic. As for the central government, we asked about various policies including the supply of vaccines. For example, we asked respondents what they thought of some descriptions such as “Vaccinations are progressing well across the country.” and “The government’s response to the pandemic has been successful.” with a 5-point Likert scale ranging from 1 for “agree” to 5 for “disagree”. Similar questions were used for the municipal government. We use descriptions such as “Vaccinations are progressing well in your municipality.” and “Your municipality has adequate measures to prevent COVID-19 infection.” ^15^

#### Health behaviors and mental health

Vaccination may change people’s awareness of COVID-19 and their daily behaviors. We asked about the compliance of daily infection prevention behaviors such as handwashing, mask-wearing, and the frequency of high-risk behaviors such as attending large group events and eating at restaurants. From these questions, total scores on infection preventive behaviors and high-risk behaviors were calculated. We also used Kessler Psychological Distress Scale (K6) to evaluate the mental health of respondents (Kessler et al., 2003). Detailed definitions and descriptive statistics on each item are reported in Appendix Table A1.

### 4.4 Variables for Subsamples Analysis

#### Chronic diseases

According to the Centers for Disease Control and Prevention (CDC), people with certain medical conditions are more likely to get severely ill from COVID-19 (CDC, 2022). To obtain information on the health status, we asked whether respondents have the following chronic diseases: hypertension; diabetes; bronchitis/pneumonia; angina; myocardial infarction; stroke (cerebral infarction or cerebral hemorrhage); chronic obstructive pulmonary disease (COPD); cancer; and chronic pain. Descriptive statistics on each item are reported in Appendix Table A2.

#### Interpersonal Trust

To measure interpersonal trust, many previous studies have used people’s trust in their neighbors (Rotter, 1967; Yamagishi, 1988; Siegrist and Bearth, 2021). In this study, we use a scale proposed by Siegrist and Bearth (2021). The following four items were used to measure respondents’ interpersonal trust: 1) “If given the chance, most people in your city would try to take advantage of you”; 2) “Most people in your city are too busy looking out for themselves to be helpful”; 3)“Most people in your city are honest”; 4)“Most people in your city tell a lie when they can benefit from doing so”.^16^ The respondents could indicate their agreement with the statements, using the scale of 1 (completely agree) to 7 (do not agree at all). The total score of these items is used as an index of interpersonal trust. To align the scale direction of the items, we reversed the scale for the third questionnaire item.^17^ The total score ranges from 4 to 28, and a higher value indicates a higher level of interpersonal trust.

#### Socioeconomic status

It is well known that people’s political opinions differ according to their SES. We use educational background and income as indicators of SES. We used whether a person has a college degree as an indicator of educational background. For income, we asked respondents what their annual household income would have been in 2020. We also allowed for responses such as “I don’t know” or “I don’t want to answer.”

### 4.5 Descriptive Statistics

Table 1 reports descriptive statistics for all respondents, people who are eligible for priority vaccination (PV) for older people (over 65 years), and those who are not eligible for PV (under 65 years). Our online survey received responses from a total of 30,892 people, of which 13,852 were eligible for PV and 17,040 were not eligible for PV.

**Table 1:** Descriptive Statistics

|  | All respondents |  | Eligible for PV<br>(Age 65–67) |  | Not eligible for PV<br>(Age 62–64) |  |
| --- | --- | --- | --- | --- | --- | --- |
|  | Mean | SD | Mean | SD | Mean | SD |
| Vaccination rate (%) | 66.7 | 0.47 | 86.9 | 0.34 | 50.3 | 0.50 |
| Individual characteristics: |  |  |  |  |  |  |
| Woman (%) | 23.0 | 0.42 | 21.6 | 0.41 | 24.1 | 0.43 |
| College graduate (%) | 59.2 | 0.49 | 59.5 | 0.49 | 58.9 | 0.48 |
| Above median income | 62.0 | 0.49 | 58.4 | 0.49 | 64.8 | 0.48 |
| Having jobs (%) | 64.4 | 0.48 | 54.3 | 0.50 | 72.5 | 0.45 |
| Having chronic diseases (%) | 44.3 | 0.47 | 47.0 | 0.50 | 42.2 | 0.49 |
| Living in high-infection area (%) | 55.0 | 0.50 | 54.8 | 0.50 | 55.0 | 0.50 |
| Subjective efficacy of COVID-19 vaccine (%) | 79.6 | 26.35 | 80.5 | 25.96 | 78.9 | 26.65 |
| Interpersonal trust | 18.0 | 3.16 | 18.0 | 3.18 | 18.0 | 3.13 |
| Trust on scientists | 6.3 | 2.36 | 6.3 | 2.34 | 6.3 | 2.37 |
| Relative Risk Aversion | 0.2 | 0.003 | 0.2 | 0.003 | 0.2 | 0.003 |
| Political opinion (1:Agree – 5:Disagree): |  |  |  |  |  |  |
| (a) Vaccinations are progressing well in your municipality. | 2.855 | 1.11 | 2.802 | 1.09 | 2.899 | 1.21 |
| (b) Your municipality has adequate measures to prevent COVID-19 infection. | 3.098 | 0.92 | 3.093 | 0.93 | 3.102 | 0.92 |
| (c) Vaccinations are progressing well across the country. | 3.554 | 1.07 | 3.565 | 1.07 | 3.545 | 1.08 |
| (d) The government’s response to the pandemic has been successful. | 3.991 | 1.08 | 4.007 | 1.08 | 3.978 | 1.07 |
| Observation | 30,892 |  | 13,852 |  | 17,040 |  |

Because of the age-based priority rule, COVID-19 vaccination rates vary widely between people aged 65 and older and those under 65. We use the fraction of people who completed two doses as vaccination rates. While vaccination rates for people under 65 is a little over 50%, it reaches nearly 87% for those aged 65 and older. As a result, the difference in vaccination rate between the two groups amounts to over 36%.

The fraction of women is 23% because more men than women were registered for the survey. The fraction of college graduates is nearly 60%, and there is no significant difference between the two groups. The employment rate and annual income of people under 65 years are higher than that of people aged 65 years and older. This is probably because more people retire at an older age. In addition, 45% of the respondents have chronic diseases listed in Section 4.4, and older people are more likely to have those. The subjective efficacy of the COVID-19 vaccine, interpersonal trust, and trust in scientists do not differ between the two groups.^18^ Regarding subjective efficacy of the COVID-19 vaccine, respondents of our survey believe that the COVID-19 vaccine prevents 79.6 % of fully vaccinated people from developing severe conditions.^19^

Descriptive statistics for political opinions are the mean value of a 5-point Likert scale ranging from 1 for “agree” to 5 for “disagree” for each political description. Descriptions (a) and (b) are related to municipal government policies against the pandemic, and descriptions (c) and (d) represent ones for the central government. Based on the mean value, there does not appear to be a significant difference in political opinions between the two groups. Although the mean values suggest that PV eligibility has little effect on respondents’ political opinions, the simple comparison is affected by various confounding factors. In the following section, we describe how to identify the effect of eligibility of PV and vaccination status on political opinions using regression discontinuity design.

## 5 Empirical Strategy

### 5.1 Identification

We identify the political impact of COVID-19 vaccination by exploiting the age-based priority policy. The Japanese government has made it possible for people born before April 1, 1957, who have reached or will reach the age of 65 during the fiscal year 2021, to be vaccinated before younger people. We use year-month of birth to determine if the respondent is eligible for priority vaccination (PV). For example, people born in March 1957 are eligible for PV, whereas people born in April 1957, whose birthdays are one month later, are not. For simplicity, we will refer to the year-month birth threshold as the “age 65 threshold”. The age 65 threshold influences the probability of vaccination because of PV eligibility.

The age-based priority is useful to identify the political impact of vaccination while excluding confounding factors. It is noteworthy that the age 65 threshold represents whether a person will *turn 65 within the fiscal year 2021*. Some social security systems in Japan also use age 65 as a threshold for their eligibility. However, the timing of the change in these eligibilities is *when a person turns 65*. For example, the eligibility for some pensions and public long-term care insurance changes when they turn 65.^20^ Hence, the age 65 threshold for vaccination differs from the thresholds for other systems and excludes the political impact other than vaccination.

We use whether the person is older than the age 65 threshold as an instrumental variable (IV) for her vaccination status. Because our running variable is age-in-month, it is unlikely that we have a general manipulation problem around the threshold. The institutional feature explained above implies that the age 65 threshold should affect outcomes only through vaccination status, which is a desirable property of IV. We will statistically check the validity of identification in Section 6.1. The COVID-19 vaccine should be given two doses, three to four weeks apart.

### 5.2 Estimation

Our estimation strategy relies on the fuzzy RD design using the eligibility for PV as an instrument for vaccination status. In our baseline specification, the effect of COVID-19 vaccination on political support and other outcomes is expressed as:

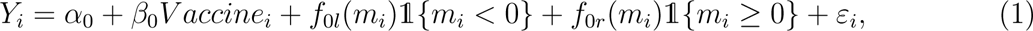

where *Y_i_* is the outcome variable for political opinions or other responses of respondent *i*. The variable *Vaccine_i_* stands for a dummy variable which is equal to 1 if the respondent *i* has received two doses of the COVID-19 vaccine. The variable *m_i_* represents age-in-months and is normalized to zero at the age 65 threshold. Therefore, the respondent *i* is eligible for PV for elderly people if *m_i_ ≥* 0. We use *m_i_* as a running variable in this estimation. The functions ƒ_0_*_l_*(*·*) and ƒ_0_*_r_* (*·*) respectively signify the function of the running variable *m_i_* on the left and right side of the threshold. The parameter of interest is *β*_0_, denoting the effect of COVID-19 vaccination on political opinions and other outcomes.

The first-stage regression is

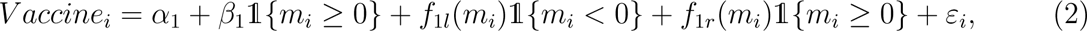

where a dummy variable 1{*m_i_ ≥* 0} takes a value 1 if *m_i_ ≥* 0 and respondent *i* is eligible for PV. The dummy variable is used as an instrument for vaccination status. The function ƒ_1_*_l_*(*·*) and ƒ_1_*_r_* (*·*) respectively signify the function of the running variable *m_i_* on the left and right side of the threshold. For estimation, we adopt nonparametric local linear and local quadratic regressions with robust confidence intervals proposed by Calonico et al. (2014)

Another important parameter is the political impact of the eligibility of PV. The reduced-form specification of the above model is

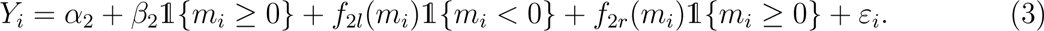

The parameter *β*_2_ represents the impact of eligibility for PV on political opinions and other outcomes. This parameter corresponds to the one from a sharp RD, which is equivalent to the discontinuity at the age 65 threshold in each figure of outcomes.

The above specifications do not control covariates *X_i_* because the RD design can identify the parameter of interest without controlling those variables if the continuity assumption holds. We check the continuity assumption of predetermined characteristics in the following section.

The advantage of this specification is that respondents with missing covariates can be included in the estimation. About 20% of respondents in our survey did not provide income information, but these respondents can also be used for estimation. As a robustness check, we also report estimation results when we control for various covariates such as gender, income, employment, education level, health status, and living areas, but this estimation comes at the expense of sample size.

## 6 Empirical Results

### 6.1 Continuity Tests

As a validity check for RD design, we examine the continuity of various predetermined characteristics at the age 65 threshold. The definitions of the ten variables for predetermined characteristics are given in Appendix Table A3. Appendix Table A4 reports estimates for the continuity test, and we find that almost all characteristics do not significantly change at the threshold. These results strongly support the continuity assumption of our RD design. The only exception is the change in the fraction of college graduates at the 5% significance level. However, there is no institutional reason for the college graduation rate to change at the threshold and this may reflect the noise of our data. We find an irregular value at the threshold on this variable, which may drive the result. To check this possibility, we show the results of donut-hole RD (Barreca et al., 2016) by excluding respondents who were born just one month before and after April 1, 1957. Estimates for the donut-hole RD indicate no statistically significant jump, including college graduates. These results show that there are no significant discontinuities in key characteristics at the threshold.^21^

### 6.2 Complier Characteristics

The IV estimator identifies the local average treatment effect (LATE), which is the effect of COVID-19 vaccination on the “complier” subpopulation. In our context, compliers are people who receive vaccination only when they are eligible for PV. Before moving to the main results, we check the complier characteristics to clarify which people our estimator focuses on.

To do this, we adopt a method proposed by Marbach and Hangartner (2020) and plot the characteristics of each player in Appendix Figure A2.^22^ Always (never) takers are persons who (do not) take the COVID-19 vaccine regardless of their eligibility for PV. In short, compared with never-takers, compliers have more faith in the efficacy of the COVID-19 vaccine and more trust in scientists. Compliers believe that the COVID-19 vaccine prevents more than 80% of fully vaccinated persons from becoming severe conditions, but never-takers believe that it is about 66%. Also, the score for trust in scientists is 16% higher for compliers than for never-takers.^23^ These results indicate that our fuzzy RD design estimates the effect of COVID-19 vaccination among a less anti-vaccine, more science-trusting population.

### 6.3 First Stage Results

Next, we examine how the eligibility of PV affects vaccination rates. As in many other countries, receiving two doses of COVID-19 vaccination is officially recommended in Japan because the first dose is considered insufficient to develop an antibody. Figure 2 presents, for each age-in-month, the percentage of respondents who received two doses, at least one dose, and only one dose. The horizontal axis represents the age-in-month standardized to zero at the age 65 threshold. People on the right side of the threshold are eligible for PV.

**Figure 2:**
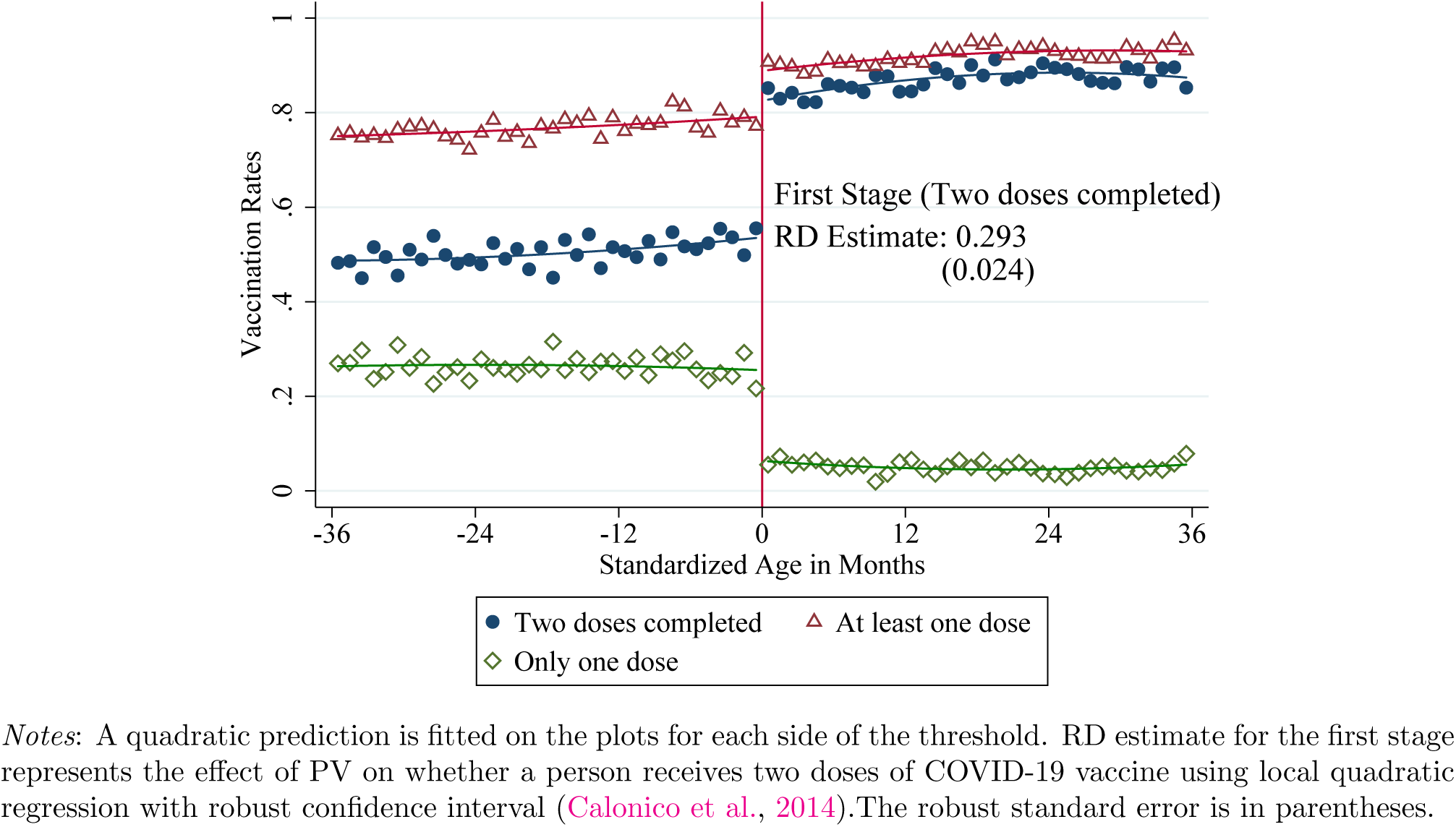
The Effect of Eligibility of Priority Vaccination on Vaccination Rate

Figure 2 shows that the eligibility of PV significantly affects vaccination rates. Among those who are not eligible for PV, about 80% have received at least one dose, but only half have completed two doses. Then the age 65 threshold changes these vaccination rates. The most obvious change is the percentage of respondents who completed two doses. The RD estimate indicates that PV eligibility increases the percentage of respondents who completed two doses by 29.3 percentage points. Compared with the mean before the cut-off age (54%), this increase amounts to more than 50 percent increase in vaccination rates for two doses. The percentage of respondents who have received at least one dose also increases at the threshold. By contrast, the percentage of those who have received only one dose decreases at the threshold, which reflects the increases in those who have received the second dose because of PV eligibility.

The first stage estimates represent the effect of eligibility of PV on the probability of completing two doses. It is important to note that our estimation strategy cannot separately identify the effect of the first dose and the effect of the second dose. This is because, as shown in Figure 2, the eligibility of PV (instrumental variable) affects both the probability of receiving the first dose and the probability of receiving the second dose. Our IV estimates for the parameter of interest in equation (1) represents the effect of COVID-19 vaccination, which is a mixture of the effect of the first dose and the effect of the second dose.

### 6.4 The Political Impact of COVID-19 Vaccination

In this section, we show estimation results for the political impact of COVID-19 vaccination. Figure 3 graphically presents the relation between the eligibility of PV and political support regarding government COVID-19 policies. Panel (a)-(b) represents the support for municipal government, and Panel (c)-(d) represents ones for the central government. Each outcome is represented with a 5-point Likert scale ranging from 1 for “agree” to 5 for “disagree”. Based on our political descriptions, the lower the outcome value, the higher the political support.

**Figure 3:**
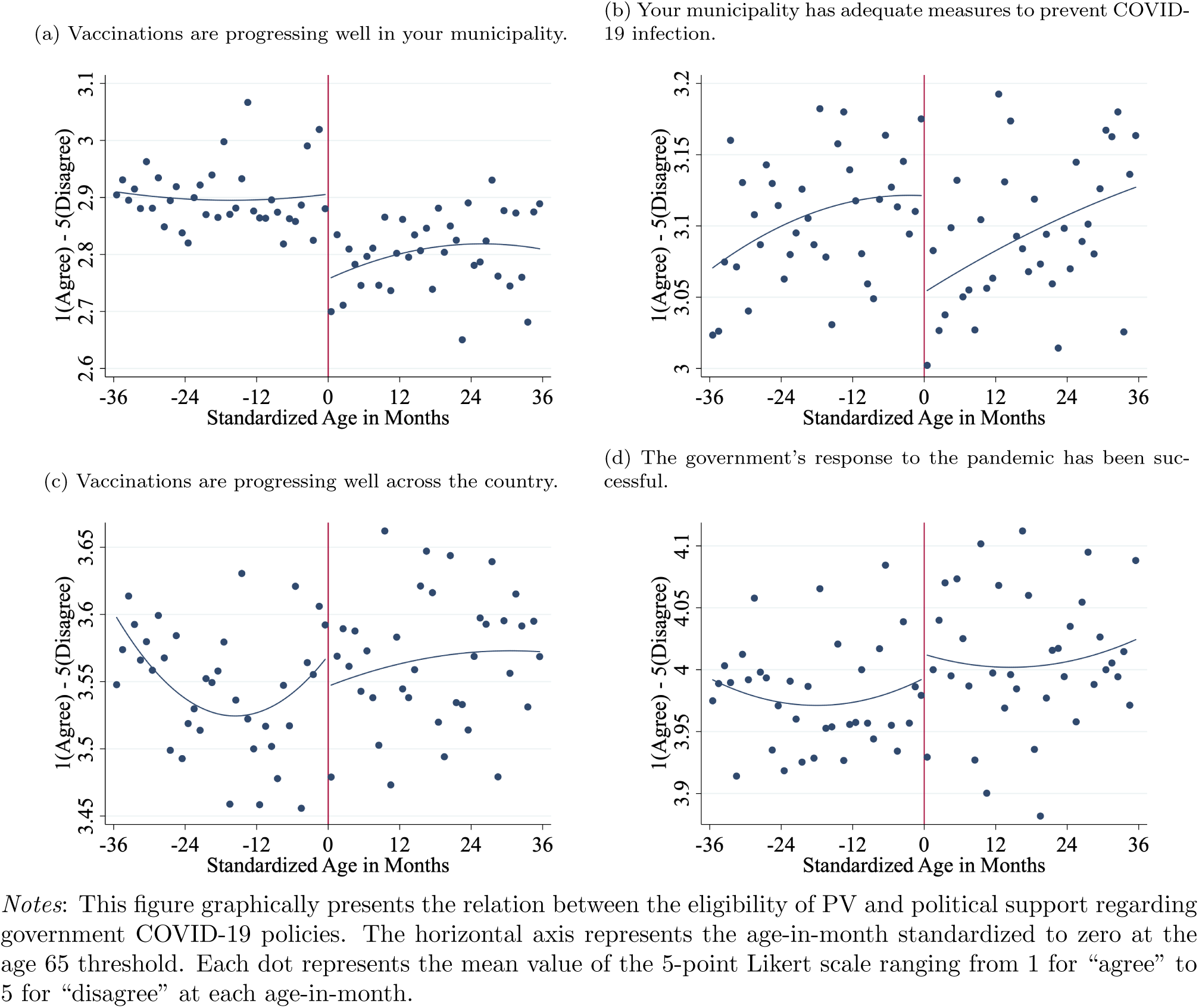
The Effect of Eligibility of Priority Vaccination on Political Support

A critical takeaway from Figure 3 is that while the eligibility of PV increases political support for infection control measures by municipalities, it does not affect support for policies by the central government. The discontinuity of outcome at the threshold in Panel (a) shows that PV eligibility increases respondents’ favorable opinions on the progress of COVID-19 vaccination in their municipalities. Panel (b) also shows that PV eligibility increases favorable opinions on infection control measures by municipalities. Meanwhile, outcomes in Panel (c) and (d) have no discontinuity at the threshold. These figures indicate that PV eligibility does not significantly impact respondents’ opinions on the progress of COVID-19 vaccination nationwide and the central government’s responses to the pandemic.

Table 2 presents estimates for the effect of COVID-19 vaccination on political opinions based on the fuzzy RD design. The first and third columns respectively present fuzzy RD estimates from the local linear regressions and the local quadratic regression using the robust confidence intervals proposed by Calonico et al. (2014). The descriptions (a)-(d) correspond to the ones in Figure 3 Panel (a)-(d). Estimates for description (a) indicate that COVID-19 vaccination increases favorable opinions on vaccination progress in a municipality by 0.65-0.73 points and these are statistically significant (note that negative estimates imply increases in political support). These estimates are equivalent to 22.8–25.4% of the mean outcome value. Similarly, estimates for description (b) indicate that COVID-19 vaccination increases favorable opinions on municipal government’s infection control measures by 0.41–0.45 points, and these are also statistically significant. These estimates are equivalent to 13.1–14.6% of the mean of outcome value. In contrast, as presented in Figure 3, COVID-19 vaccination does not significantly affect opinions on policies by the central government. The fact that estimates for descriptions (c) and (d) are not statistically significant implies that opinions on the progress of vaccination nationwide and the central government’s responses to the pandemic are largely unaffected by one’s COVID-19 vaccination status.

**Table 2:**
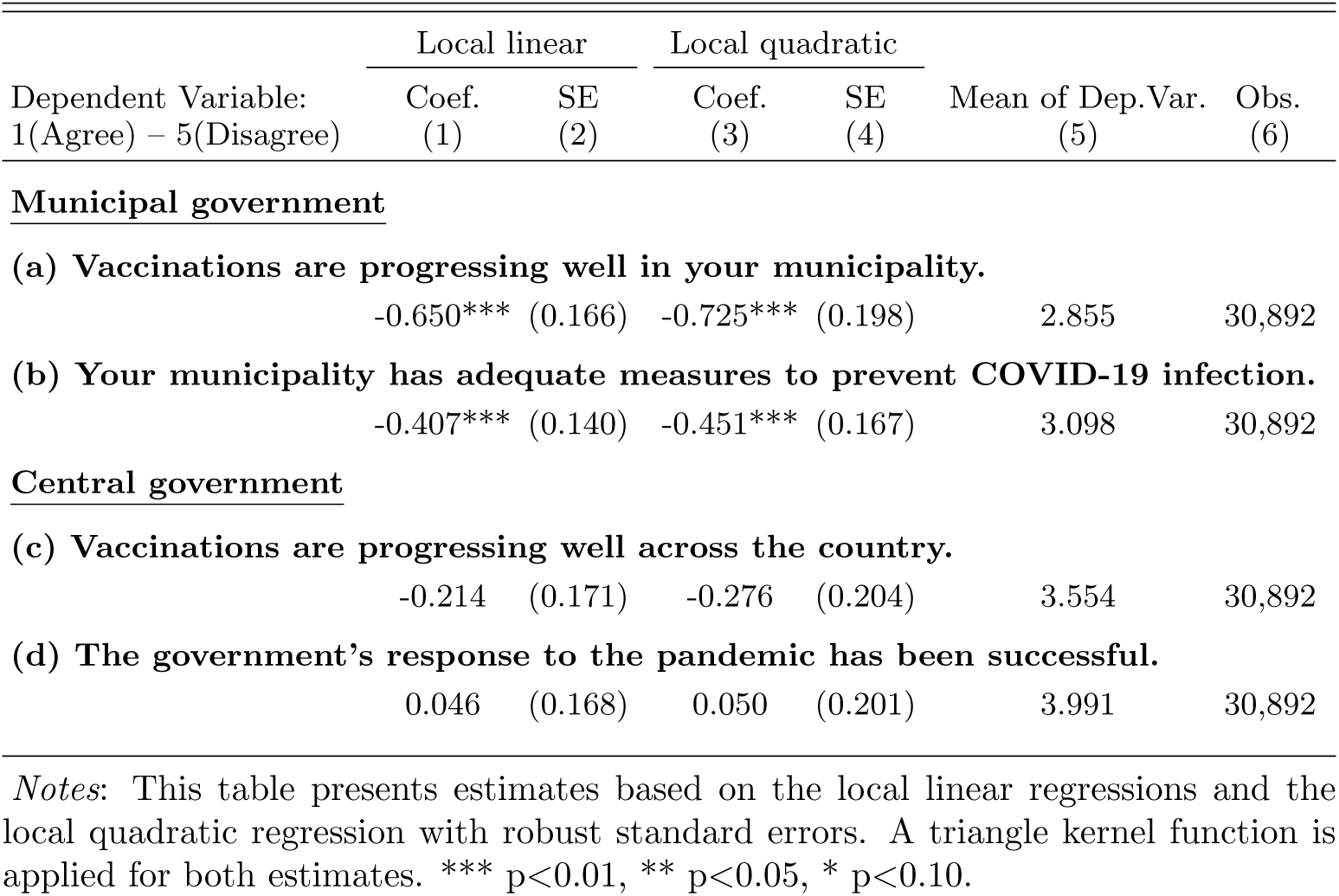
Estimates for the Political Impact of COVID-19 Vaccination

The above results suggest that the COVID-19 vaccination may have made the efforts of municipal governments to deal with the pandemic more visible. Each municipal government is responsible for administering the COVID-19 vaccination program (securing medical personnel, preparing vaccination sites, distributing vaccination coupons, etc.). Receiving a vaccination coupon and getting vaccinated at a vaccination site might make the municipal government’s effort more visible and serve as important political information about their policies against the pandemic. Although the central government is responsible for securing COVID-19 vaccines and delivering them to each municipality, it is not directly involved in the actual implementation of vaccination. The estimation results that COVID-19 vaccination does not influence opinions on the central government policies may indicate that vaccination does not make the central government’s efforts visible.

### 6.5 COVID-19 Vaccination and Reciprocity

#### 6.5.1 Health Risks

In the following section, we explore factors that drive the impact of COVID-19 vaccination on the political support for the municipal government. First, we examine the influence of people’s health risks. It is widely known through government and media publicity that people with underlying medical conditions are more likely to become critically ill from COVID-19. The upper panel of Figure 4 presents estimates for people with and without any of the chronic diseases listed in Section 4.4. Comparing these estimates, people with chronic diseases are more likely to increase favorable opinions on vaccination progress and the adequacy of infection control measures in their municipality through COVID-19 vaccination. These estimates may reflect that people with chronic diseases are more likely to be grateful for vaccination and take it as important political information. Therefore, the greater impacts on people with chronic diseases may indicate the importance of reciprocal motivation in political support for the municipal government.

**Figure 4:**
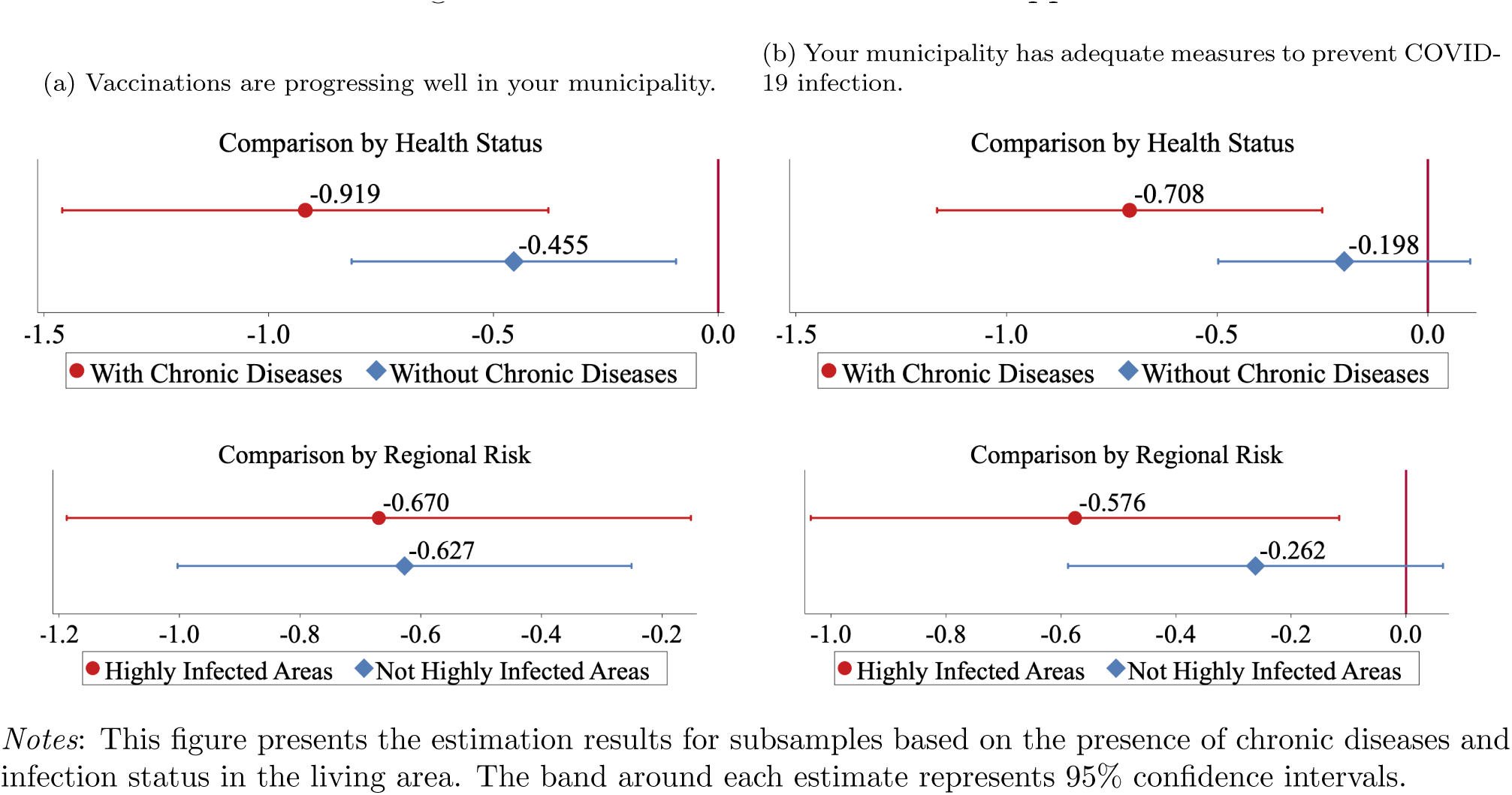
Health Risks and Political Support

The large variation in estimates by health risk with COVID-19 suggests that there may be regional differences in the political impact of COVID-19 vaccination. As a general characteristic of infectious diseases, the risk of COVID-19 infection is higher in large, densely populated areas. We divide respondents’ living areas into highly infected areas and others at the prefecture level and separately estimate the political impact of COVID-19 vaccination.^24^ The lower panel of Figure 4 compares estimates for the two areas. The estimation results indicate that people living in highly infected areas are more likely to increase favorable opinions of infection control measures by municipalities. However, the impact on opinions of vaccination progress is almost identical between the two groups. Therefore, in terms of risk from COVID-19, an individual’s health condition was a more important factor in the political impact of vaccination than the risk of infection in living areas.

#### 6.5.2 Interpersonal Trust

Given the positive reinforcement between interpersonal trust and reciprocity (Ostrom and Walker, 2003) and empirical evidence on their positive correlation (Altmann et al., 2008; Dohmen et al., 2008), people with high interpersonal trust might increase their political support for the government through reciprocal motives.

To investigate the relationship between interpersonal trust and political support, we split the sample based on the median of the interpersonal trust score and estimate using each sample. We also report the results on the persons with chronic diseases because the political impact of vaccination is much larger among this subsample. The results are summarized in Figure 5. In this figure, the lower (higher) score of the interpersonal trust index represents the lower (higher) level of interpersonal trust. Panels (a) and (b) in Figure 5 show a clear difference of the political impact of vaccination by the level of interpersonal trust, especially among persons with chronic diseases. In the persons with the score below the median, the point estimate on the impact of vaccination among persons with chronic diseases is very close to zero (−0.178) in Panel (a), supporting the null effects of vaccination among persons with the low level of interpersonal trust. However, estimates increase as the level of trust increases: Among the persons with the high score, the estimate is -2.005 which amounts to 63 % of the standard deviation of the interpersonal trust index. In Panel (b), we also find a clear difference according to the level of trust. These results suggest that people might increase their political support through reciprocal motivation.

**Figure 5:**
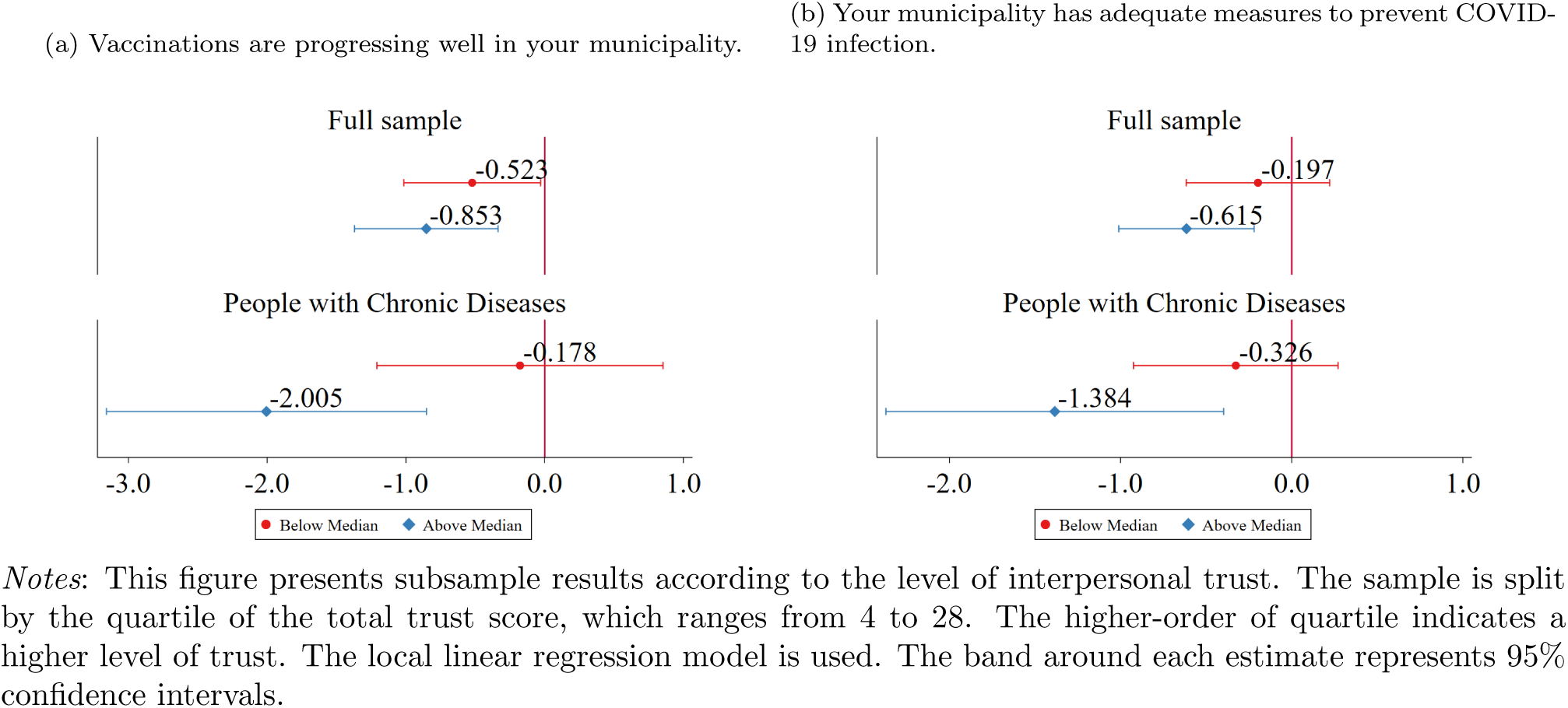
Interpersonal Trust and Political Support

### 6.6 Socioeconomic Status and Political Support

The relationship between SES and political behavior has long been a central agenda of political science. To examine which people change their political opinions because of vaccination, we estimate the political impact of COVID-19 vaccination for each SES group. We use educational background and income as main components of SES. For education, we divide respondents based on whether they had a college degree. For income, we divide respondents according to whether their household income is above the median among the respondents. We also investigate whether the political impact of COVID-19 vaccination varies by gender.

Figure 6 summarizes the local linear estimates by SES and gender. With this estimation, we find a substantial difference in the political impact of COVID-19 vaccination by SES. Most importantly, people with low SES are more likely to increase political support because of their vaccination uptake. Estimates in Panel (a) indicate that those without a college degree and those with lower incomes are more likely to have a favorable opinion of vaccination progress in their municipality. The relation between SES and the political impact of vaccination becomes even more pronounced for infection control measures by municipalities. Estimates in Panel (b) show that COVID-19 vaccination significantly increases favorable opinions of infection control measures by municipalities among those without college degrees and those with low incomes. By contrast, estimates for those with college degrees and those with high incomes are statistically insignificant.

**Figure 6:**
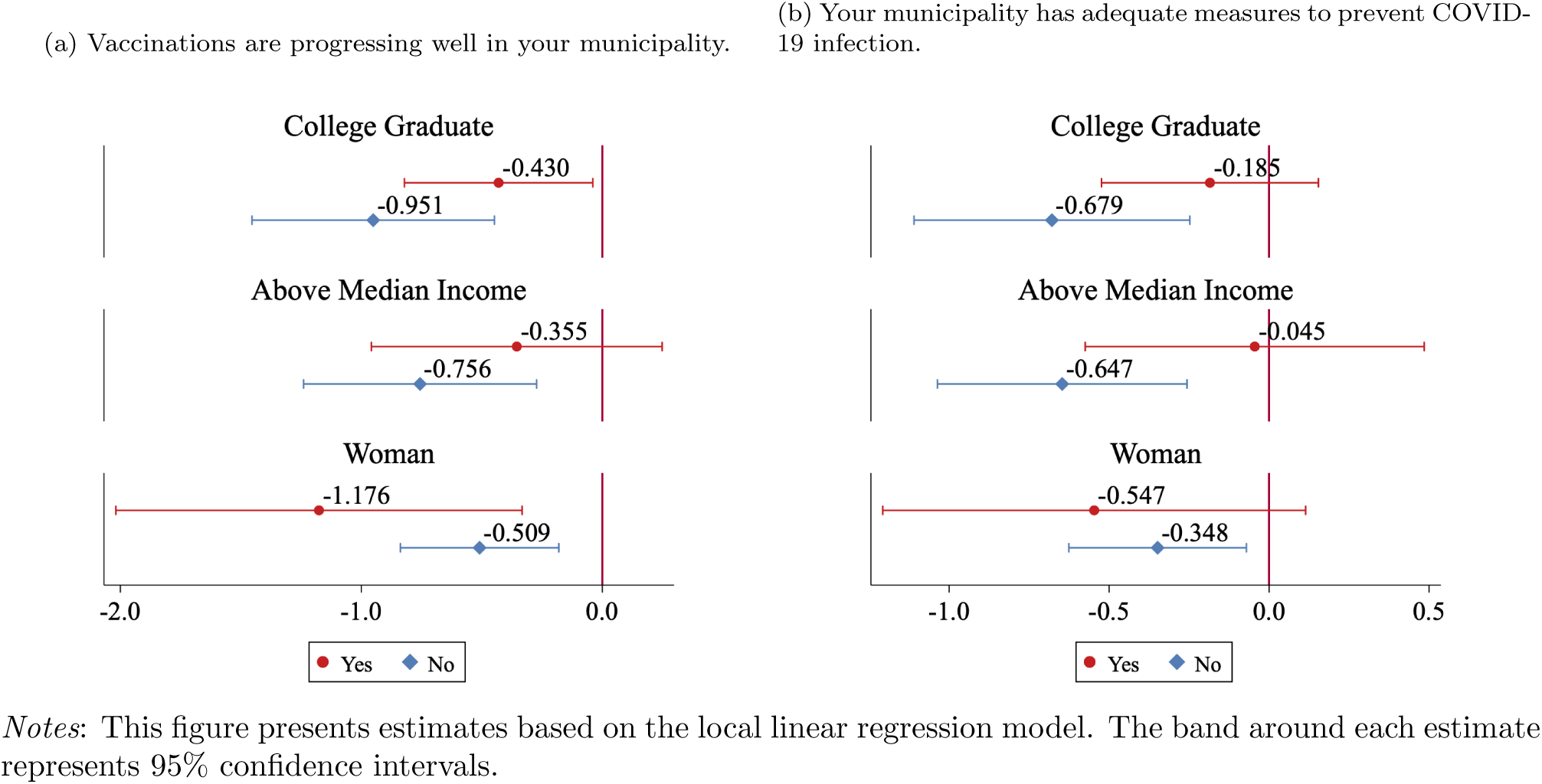
SES and Political Support

The above estimates indicate that when evaluating government policies, people with low SES are more likely to overvalue their personal experiences than those with high SES. The vaccination status of a single person is supposed to provide little information on the overall vaccination progress and the adequacy of infection control measures. However, if people have limited knowledge about policy consequences in advance, they may evaluate government policies based on their personal experiences, such as having been vaccinated. Angelucci and Prat (2021) empirically show that people with low SES tend to have less accurate information about political news than those with high SES. Our findings can be interpreted as a result of information inequality about government policies by SES: Estimates in Figure 6 suggest that people with lower SES have limited prior political knowledge and that COVID-19 vaccination significantly updates their political opinions. By contrast, those with higher SES may have had sufficient prior political knowledge, and thus their political opinions were not affected by vaccination.

Previous studies have shown that people with low SES tend to have poorer health than those with high SES (Cutler et al., 2011). In addition, high SES individuals behave more prosocially and trust more (Andreoni et al., 2021). Therefore, our subsample analysis may have confused the two factors of the political impact of vaccination: reciprocal motive and political knowledge. For a more careful interpretation of our subsample analysis, we check the correlation between variables related to reciprocal motive and political knowledge. Table 3 presents correlation coefficients and indicates that variables related to reciprocal motive (chronic diseases, interpersonal trust) and ones related to political knowledge (college graduate, income) are not strongly correlated in our sample. Thus, it can be argued that both reciprocal motive and political knowledge are important factors of the political impact of vaccination.

**Table 3:** Correlation between variables

|  | College Graduate | Above Median Income |
| --- | --- | --- |
| With Chronic Diseases | 0.040 | 0.012 |
| Interpersonal Trust Score | 0.029 | 0.020 |

Notes: This table presents correlation coefficients between variables.

Another important finding is that the political impact of COVID-19 vaccination varies by gender. Estimates in Panel (a) show that women are more likely than men to increase their favorable opinion of vaccination progress in their municipality. The gender difference can be attributed to several reasons. First, women may be less informed about the government policies in advance. Existing studies show that women are typically less knowledgeable about politics and government than their male counterparts (Lizotte and Sidman, 2009). Therefore, women might be more likely to update their political opinions because of vaccination. Second, it is also possible that women expressed more gratitude to the government for vaccinations because women are generally more reciprocal than men (Croson and Buchan, 1999).

## 7 Discussion

### 7.1 Vaccine, Health Behaviors, and Mental Health

There are other possible mechanisms for the impact of COVID-19 vaccination on political support. One hypothesis is that vaccination may have improved people’s quality of life (QOL), which may have increased their political support. We examine whether COVID-19 vaccination affects people’s lifestyles and mental health to test this hypothesis. During the pandemic, people have been constrained in social interactions to prevent infection. For example, people were forced to refrain from eating and drinking in restaurants and participating in large-scale events due to the high risk of infection. The easing of behavioral restrictions due to vaccination may improve people’s QOL and lead to their political support for the government. It is also possible that vaccination reduces the fear of COVID-19 and relieves psychological stress (Agrawal et al., 2021), which may contribute to increased political support.

Table 4 presents estimates for the impact of COVID-19 vaccination on lifestyles and mental health. We examine people’s lifestyles in terms of infection prevention and high-risk behaviors. Both behaviors are quantified by the total score of the responses to questionnaire items summarized in Appendix Table A1.^25^ For example, infection prevention behaviors include how careful they are about washing hands and wearing masks. High-risk behaviors, on the other hand, include how often they visit restaurants and participate in large-scale events.

**Table 4:** The Impact of COVID-19 Vaccination on Health Behaviors and Mental Health

| Dependent Variable | Local linear |  | Local quadratic |  | Mean of Dep.Var.<br>(5) | Obs.<br>(6) |
| --- | --- | --- | --- | --- | --- | --- |
|  | Coef.<br>(1) | SE<br>(2) | Coef.<br>(3) | SE<br>(4) |  |  |
| **<u>Health behavior</u>** |  |  |  |  |  |  |
| **(a) Infection prevention behaviors** (High scores indicate low prevention awareness.) |  |  |  |  |  |  |
| All respondents | 0.513 | (1.067) | 0.540 | (1.250) | 23.893 | 30,892 |
| With chronic diseases | 2.154 | (2.321) | 2.312 | (2.703) | 24.156 | 13,693 |
| Without chronic diseases | 0.162 | (1.384) | 0.224 | (1.598) | 23.682 | 17,199 |
| **(b) High-risk behaviors for infection** (Higher score indicates more high-risk behavior.) |  |  |  |  |  |  |
| All respondents | -0.188 | (0.786) | -0.257 | (0.920) | 24.800 | 30,892 |
| With chronic diseases | 0.617 | (1.346) | 0.563 | (1.564) | 24.958 | 13,693 |
| Without chronic diseases | -0.729 | (0.901) | -0.900 | (1.056) | 24.673 | 17,199 |
| **<u>Mental health</u>** |  |  |  |  |  |  |
| **(c) Fear of COVID-19** (High scores indicate high fear of COVID-19.) |  |  |  |  |  |  |
| All respondents | -0.526 | (1.142) | -0.337 | (1.341) | 24.152 | 30,892 |
| With chronic diseases | -1.714 | (1.766) | -1.746 | (2.072) | 24.376 | 13,693 |
| Without chronic diseases | -0.126 | (1.280) | -0.144 | (1.493) | 23.974 | 17,199 |
| **(d) Total score of K-6 distress scales** (High scores indicate less mental stress) |  |  |  |  |  |  |
| All respondents | 0.675 | (0.842) | 0.832 | (0.992) | 26.053 | 30,892 |
| With chronic diseases | -1.085 | (1.459) | -1.068 | (1.705) | 25.898 | 13,693 |
| Without chronic diseases | 1.777* | (0.940) | 2.182* | (1.120) | 26.176 | 17,199 |

*Notes:* This table presents estimates based on the local linear regressions and the local quadratic regression with robust standard errors. A triangle kernel function is applied for both estimates. A list of chronic diseases is summarised in Appendix Table A2. \*\*\* $p < 0.01$ , \*\* $p < 0.05$ , \* $p < 0.10$ .

Estimates in Table 4 indicate that COVID-19 vaccination does not significantly affect both infection prevention and high-risk behaviors regardless of respondents’ health condition. These estimates seem counterintuitive but could be explained by how the infection situation worsened during our survey period. As shown in Figure 1, our survey period coincided with the period of the sharpest increase in the number of infected people in Japan since the beginning of the pandemic. People’s vigilance against COVID-19 was at its peak during this period, as many patients were infected with COVID-19 and became severely ill but could not be hospitalized because of the overwhelmed health care system. As a result, people may not have relaxed their vigilance even though they had been vaccinated. Another factor that made it difficult to take high-risk behaviors was the declaration of the state of emergency. The state of emergency restricted the business hours of restaurants and other establishments, and large-scale events were refrained from being held. COVID-19 vaccination may not have increased high-risk behaviors because these behaviors were restricted regardless of vaccination status.

Our estimates also indicate that COVID-19 vaccination does not significantly affect mental health. According to estimates for all respondents, vaccination slightly reduces the fear of COVID-19 and improves mental health, but these estimates are not statistically significant. A possible reason is that the spread of the infection during the survey period continued to restrict people’s activities, and the vaccination did not sufficiently alleviate the psychological stress.

The results above, together with those in Section 6, imply that people consider the fact that they were able to get vaccinated to be important. Please remember that COVID-19 vaccines are given to those who want to receive them in the first place. Also, as discussed in section 6.2, compliers believe in the high efficacy of vaccines and have a high level of trust in science. Because of these facts, people should have a favorable perception of receiving the vaccine, even if it did not significantly improve their QOL or mental health. Therefore, the political impact of COVID-19 vaccination is mainly because people could get the vaccine, rather than changes in lifestyle or mental health.

### 7.2 Comparisons by Risk Preferences

The political impact of COVID-19 vaccination can also depend on people’s risk preferences. Risk-averse people may take the vaccination as more important political information because they want to reduce their risk of infection with COVID-19 compared to those who are not. In Section 6.4, we show that people at high risk of severe disease due to COVID-19 infection (i.e., those with chronic diseases) are more likely to increase their political support because of vaccination. In addition to actual health risks, how people perceive risk may also influence the significance of the political impact of COVID-19 vaccination.

To examine this hypothesis, we use respondents’ risk preferences measured by the Arrow-Prat absolute risk aversion. Following the existing literature, our survey elicit risk preferences using a hypothetical lottery.^26^ Specifically, respondents are asked how much they would be willing to pay for a hypothetical lottery where there is a 50% chance of winning 100,000 JPY (1,000 USD) and a 50% chance of receiving nothing. Respondents are presented with eight prices in ascending order, from 10 JPY (0.1 USD) to 50,000 JPY (500 USD). For each price, respondents are required to choose whether they buy a lottery or not. Therefore, the reservation price for the lottery should be between the price at which the respondent switches from “buy” to “do not buy”. We define the reservation price as the midpoint of the two prices. The Arrow-Prat measure of absolute risk aversion is calculated based on the reservation price.^27^

We divided the respondents into two groups based on whether their absolute risk aversion is higher than the median among respondents or not and compared the estimates. Figure 7 presents estimates of the two groups. The differences between the two groups are evident in their views on the vaccination progress in the municipality, as shown in Panel (a): Receiving COVID-19 vaccination, only people with high absolute risk aversion significantly increase favorable opinions of vaccination progress. In contrast, Panel (b) shows that the effect of COVID-19 vaccination on views on infection control measures by municipalities is almost the same between the two groups. These results indicate that not only actual health risks (chronic diseases) but also how people perceive risk (risk preferences) are possible factors in the political impact of COVID-19 vaccination, especially for vaccination progress.

**Figure 7:**
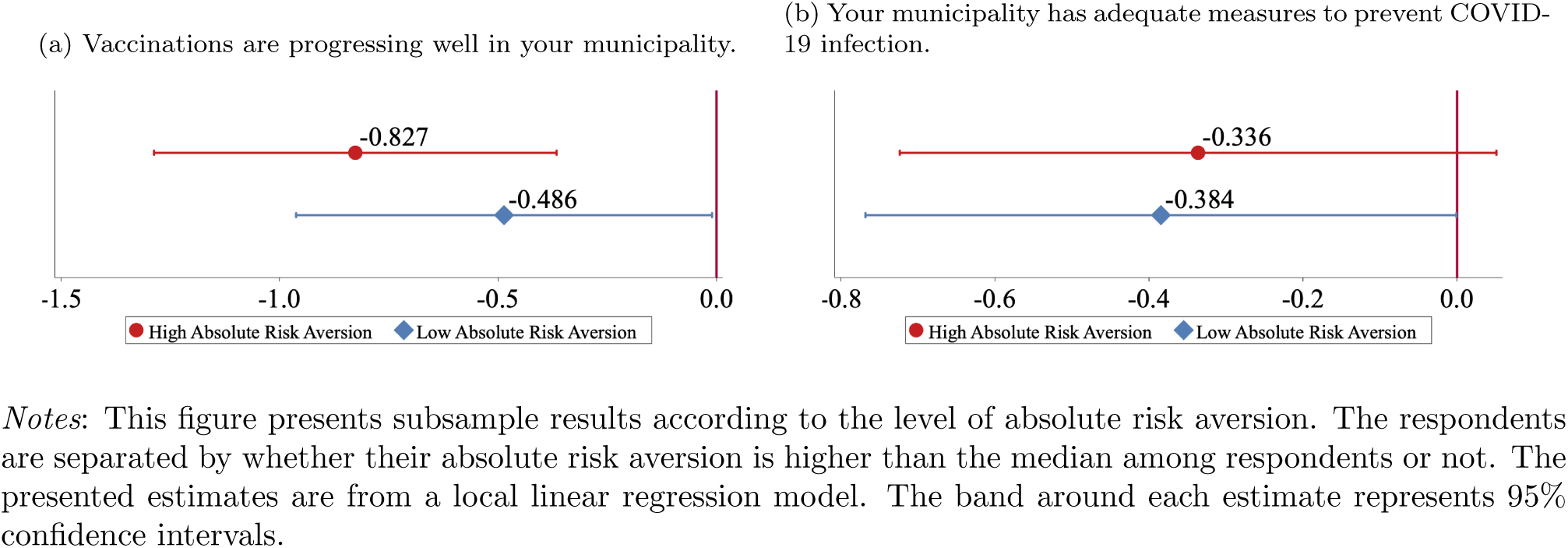
Comparisons by Risk Preferences

### 7.3 Robustness Check

While our RD estimation does not control covariates, the parameter of interest can be identified because the continuity assumption holds. Nonetheless, we report estimates based on estimation controlling respondents’ characteristics as a robustness check, as is recommended by Calonico et al. (2019). We additionally control for gender, income, having a job or not, college graduates or not, having chronic diseases or not, and living in highly infected areas or not.

We also conduct placebo tests using a series of alternative age-in-month thresholds. Specifically, we use age-in-months ranging from 12 months (one year) before and after the true threshold as alternative thresholds. After changing the threshold intentionally, we implement the RD analysis based on local linear regression one by one.

The results of the specification with covariates and the placebo test are summarized in the Appendix B.1 and B.2, respectively. In short, our main results are mostly unchanged even when we control for covariates. The results of the placebo test also show that discontinuous changes in the political support at the age 65 threshold are not due to chance.

## 8 Conclusion

During the COVID-19 pandemic, various infection control measures against COVID-19 have been implemented around the world. These policies are of great interest to the public because they are closely related to people’s lives and livelihoods. Despite the great attention, we have very limited knowledge about the political consequences of the infection control measures. This paper examines the political impact of COVID-19 vaccination, which is arguably one of the most effective infection control measures. We conducted a large-scale online survey in a timely manner and implemented RD design by exploiting Japan’s age 65 threshold for priority vaccination. Our survey was conducted when the difference in vaccination rates between those aged 65 and older and those under 65 was greatest.

Our RD estimates provide some important insights into the political impact of COVID-19 vaccination. First, COVID-19 vaccination increases favorable opinions of the vaccination progress in municipalities and infection control measures of municipal governments, while there is no significant effect on policies by the central government. The sharp contrast might reflect the primary role of municipal governments in vaccination roll-out and that their efforts become visible because of vaccination.

Second, we find suggestive evidence that reciprocal motives play an important role in the political evaluation of public policies. For example, people with chronic diseases increase political support more than those without. Because the health benefits of vaccination are high for these people, vaccination may be more appreciated by those at high risk of severe disease from COVID-19 infection. In addition, people with a high level of interpersonal trust increase political support, but those with a low level of trust do not significantly change their political opinions. Given the close relationship between interpersonal trust and reciprocity (Ostrom and Walker, 2003; Altmann et al., 2008), this result also supports the importance of reciprocal motives in political support.

Third, people with low SES are more likely to be politically influenced by COVID-19 vaccination, increasing their favorable opinions of policies by municipal governments. Previous research has shown that people with low SES tend to have less political knowledge than those with high SES (Angelucci and Prat, 2021). Our estimates indicate that COVID-19 vaccination significantly updates political information among those with less political information. This result is consistent with the theoretical prediction of voters’ learning about government policies based on rational but poorly informed voters (Rogoff, 1990; Manacorda et al., 2011).

Our study focused on voters’ evaluations of the government policies, but how the government responds to voters’ evaluations can also be an important issue. Especially in a public health crisis such as the COVID-19 pandemic, governments have an incentive to make policies that the public wants to maintain support for themselves. Government policies and the public’s evaluation of them interact with each other, which determines the long-term consequences of the policies. It is beyond the scope of this study to analyze the interaction, but this is an important avenue for future research.

## Data Availability

All data produced in the present study are available upon reasonable request to the authors

## Online Appendix

## A Additional Tables and Figures

**Table A1:** Descriptive Statistics on Perception on COVID-19 and Behaviors

|  | Mean | SD | Min | Max |
| --- | --- | --- | --- | --- |
| **Perception on COVID-19 : 1(Do not agree at all)- 5(Completely Agree)** |  |  |  |  |
| COVID-19 is very scary | 3.82 | 1.04 | 1 | 5 |
| It makes me uncomfortable when I think about COVID-19 | 3.51 | 1.09 | 1 | 5 |
| I am afraid of losing lives with COVID-19 | 3.45 | 1.13 | 1 | 5 |
| I feel anxious about being infected with COVID-19 | 3.51 | 1.06 | 1 | 5 |
| I feel anxious about my family getting infected with COVID-19 | 3.62 | 1.05 | 1 | 5 |
| I'm worried that I won't infect my family and others with the coronavirus | 3.33 | 1.11 | 1 | 5 |
| I feel anxious about creating a cluster | 2.90 | 1.16 | 1 | 5 |
| **Daily Preventive Behavior : 1(Always) - 4(Never)** |  |  |  |  |
| I disinfect hands and fingers with rubbing alcohol | 1.35 | 0.59 | 1 | 4 |
| I wash hands for 15 seconds or more using soap and hand soap | 1.53 | 0.75 | 1 | 4 |
| I gargled when I got home | 1.95 | 1.06 | 1 | 4 |
| I complied cough etiquette | 1.51 | 0.88 | 1 | 4 |
| I avoid touching eyes, nose and mouth with unwashed hands | 1.84 | 0.91 | 1 | 4 |
| I disinfected items that are easily touched by humans, such as doorknobs | 2.58 | 1.02 | 1 | 4 |
| I opened the window to ventilate the room | 1.71 | 0.81 | 1 | 4 |
| I wore a mask where there were people | 1.10 | 0.41 | 1 | 4 |
| I refrain from traveling | 1.27 | 0.67 | 1 | 4 |
| I refrain from unnecessary and unurgent going out and business trips | 1.43 | 0.68 | 1 | 4 |
| I avoid talking or speaking at close range (within 1 meter) | 1.62 | 0.75 | 1 | 4 |
| I take a social distance | 1.59 | 0.72 | 1 | 4 |
| I avoid meeting people who appear to be at high risk of infection | 1.44 | 0.79 | 1 | 4 |
| I tried not to go to places where people are crowded | 1.38 | 0.63 | 1 | 4 |
| I refrained from using the restaurant inside the store | 1.59 | 0.78 | 1 | 4 |
| **Going-out Behaviors:** |  |  |  |  |
| **1(Never), 2(Once), 3(Sometimes), 4(Once and more in a week)** |  |  |  |  |
| I visited a friend's house | 1.12 | 0.41 | 1 | 4 |
| I visited a relative's house | 1.40 | 0.69 | 1 | 4 |
| I invited friends and relatives to the house | 1.23 | 0.54 | 1 | 4 |
| I went to restaurants other than bars with my friends and colleagues. | 1.19 | 0.52 | 1 | 4 |
| I went to a Japanese pub or bar with my friends and colleagues | 1.08 | 0.34 | 1 | 4 |
| I went to restaurants other than bars by myself | 1.24 | 0.63 | 1 | 4 |
| I went to a Japanese pub or bar by myself | 1.05 | 0.29 | 1 | 4 |
| I went to a nightclub | 1.02 | 0.20 | 1 | 4 |
| I went to karaoke with multiple people | 1.02 | 0.20 | 1 | 4 |
| I went to a live house | 1.02 | 0.20 | 1 | 4 |
| I participated in group sports | 1.13 | 0.50 | 1 | 4 |
| I went to watch sports | 1.05 | 0.28 | 1 | 4 |
| I went to a sports gym | 1.19 | 0.67 | 1 | 4 |
| I went for an outdoor exercise or a walk | 2.05 | 1.22 | 1 | 4 |
| I went to pachinko / pachislot. | 1.10 | 0.44 | 1 | 4 |
| I went to a club or girl's bar with entertainment | 1.02 | 0.20 | 1 | 4 |
| I went to a sex shop | 1.02 | 0.21 | 1 | 4 |
| I got on a crowded train | 1.31 | 0.77 | 1 | 4 |
| I went to a museum / cinema | 1.12 | 0.41 | 1 | 4 |
| I went to buy something other than groceries and daily necessities | 2.45 | 1.18 | 1 | 4 |

**Table A2:** Descriptive Statistics on Chronic Diseases

| Chronic diseases | Fraction (%) | SD |
| --- | --- | --- |
| High blood pressure | 34.2 | 0.47 |
| Diabetes | 11.5 | 0.32 |
| Asthma | 2.8 | 0.16 |
| Bronchiectasis, Pneumonia | 0.8 | 0.09 |
| Atopic dermatitis | 2.0 | 0.14 |
| Periodontal disease | 13.2 | 0.34 |
| Tooth decay | 5.4 | 0.23 |
| Otitis media | 0.5 | 0.07 |
| Angina pectoris | 2.0 | 0.14 |
| Myocardial infarction | 1.4 | 0.12 |
| Cerebral hemorrhage | 1.5 | 0.12 |
| Chronic obstructive pulmonary disease (COPD) | 0.5 | 0.07 |
| Cancer | 2.5 | 0.16 |
| Chronic pain that lasts more than 3 months | 4.4 | 0.20 |
| Depression | 1.6 | 0.12 |
| Mental disorders other than depression | 1.0 | 0.10 |

**Table A3:**
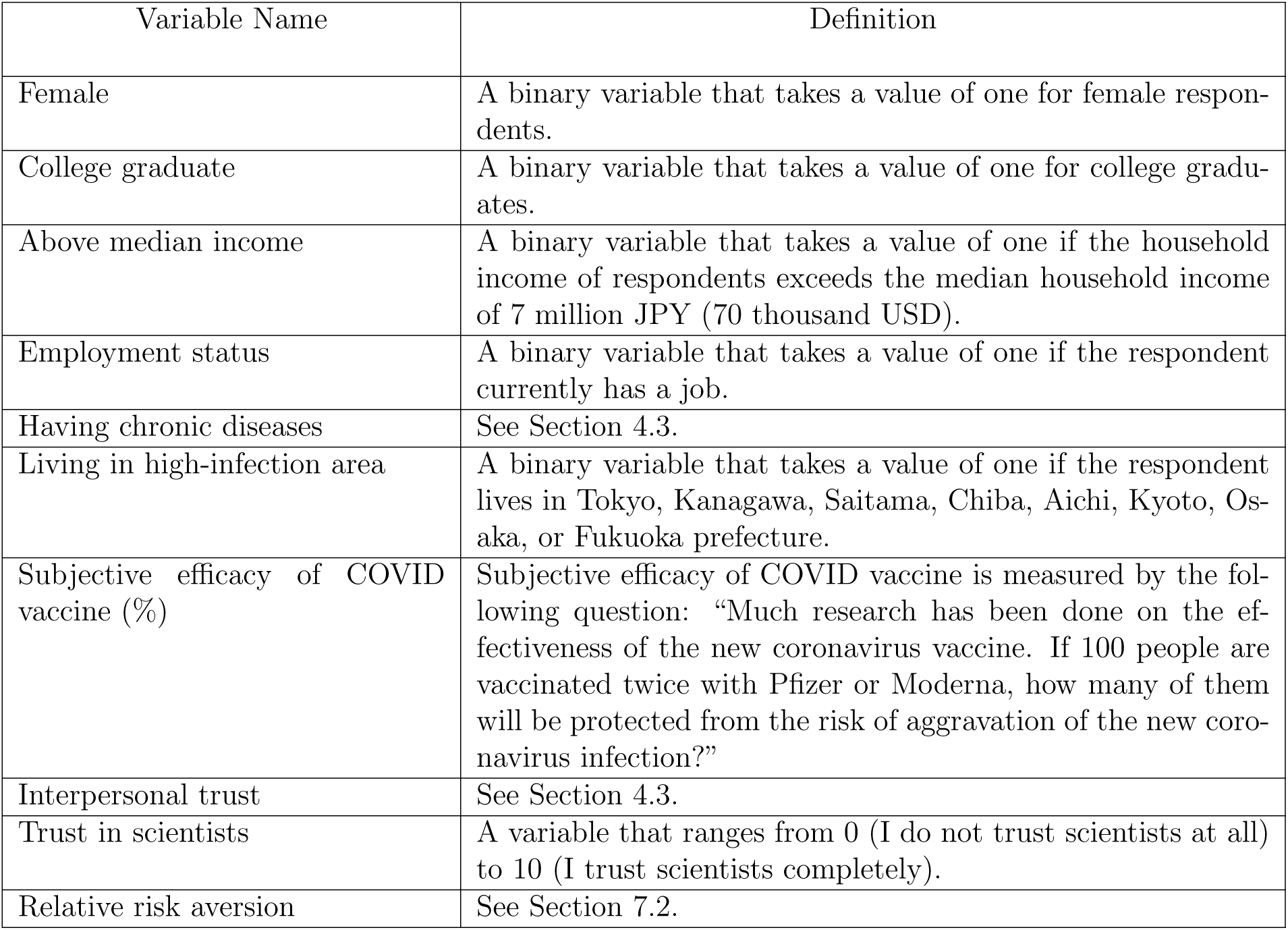
Definition of Variables on the Predetermined Characteristics

| Variable Name | Definition |
| --- | --- |
| Female | A binary variable that takes a value of one for female respondents. |
| College graduate | A binary variable that takes a value of one for college graduates. |
| Above median income | A binary variable that takes a value of one if the household income of respondents exceeds the median household income of 7 million JPY (70 thousand USD). |
| Employment status | A binary variable that takes a value of one if the respondent currently has a job. |
| Having chronic diseases | See Section 4.3. |
| Living in high-infection area | A binary variable that takes a value of one if the respondent lives in Tokyo, Kanagawa, Saitama, Chiba, Aichi, Kyoto, Osaka, or Fukuoka prefecture. |
| Subjective efficacy of COVID vaccine (%) | Subjective efficacy of COVID vaccine is measured by the following question: “Much research has been done on the effectiveness of the new coronavirus vaccine. If 100 people are vaccinated twice with Pfizer or Moderna, how many of them will be protected from the risk of aggravation of the new coronavirus infection?” |
| Interpersonal trust | See Section 4.3. |
| Trust in scientists | A variable that ranges from 0 (I do not trust scientists at all) to 10 (I trust scientists completely). |
| Relative risk aversion | See Section 7.2. |

**Table A4:** Continuity Test

|  | Mean | Full Sample |  | Donut Hole |  |
| --- | --- | --- | --- | --- | --- |
|  |  | Coef.<br>SE. | Obs. | Coef.<br>SE. | Obs. |
| Female | 0.23 | 0.03<br>(.021) | 30892 | 0.037<br>(.027) | 30005 |
| College graduate | 0.59 | -0.068**<br>(.027) | 30892 | -0.044<br>(.039) | 30005 |
| Above median income | 0.62 | -0.007<br>(.027) | 24423 | -0.041<br>(.043) | 23715 |
| Employment Status | 0.64 | -0.029<br>(.024) | 30892 | -0.031<br>(.031) | 30005 |
| Having chronic diseases | 0.45 | -0.02<br>(.023) | 30892 | 0.011<br>(.033) | 30005 |
| Living in high-infection area | 0.55 | -0.009<br>(.020) | 30892 | 0.005<br>(.025) | 30005 |
| Subjective efficacy of COVID-19 vaccine (%) | 79.62 | -0.429<br>(.996) | 30892 | -0.664<br>(1.811) | 30005 |
| Interpersonal trust | 18.05 | -0.163<br>(.115) | 30892 | -0.006<br>(.205) | 30005 |
| Trust on scientists | 6.3 | -0.027<br>(.039) | 30892 | -0.163<br>(.119) | 30005 |
| Relative Risk Aversion * 100 | 1.897 | 0.012<br>(.013) | 29372 | 0.013<br>(.016) | 28,537 |

*Notes:* : Donut hole RD excludes persons whose birthday is within just one month before and after April 1, 1957. Results from the local linear regression are reported. The score of relative risk aversion is multiplied by 100. \*\*\* p<0.01, \*\* p<0.05, \* p<0.10.

**Figure A1:**
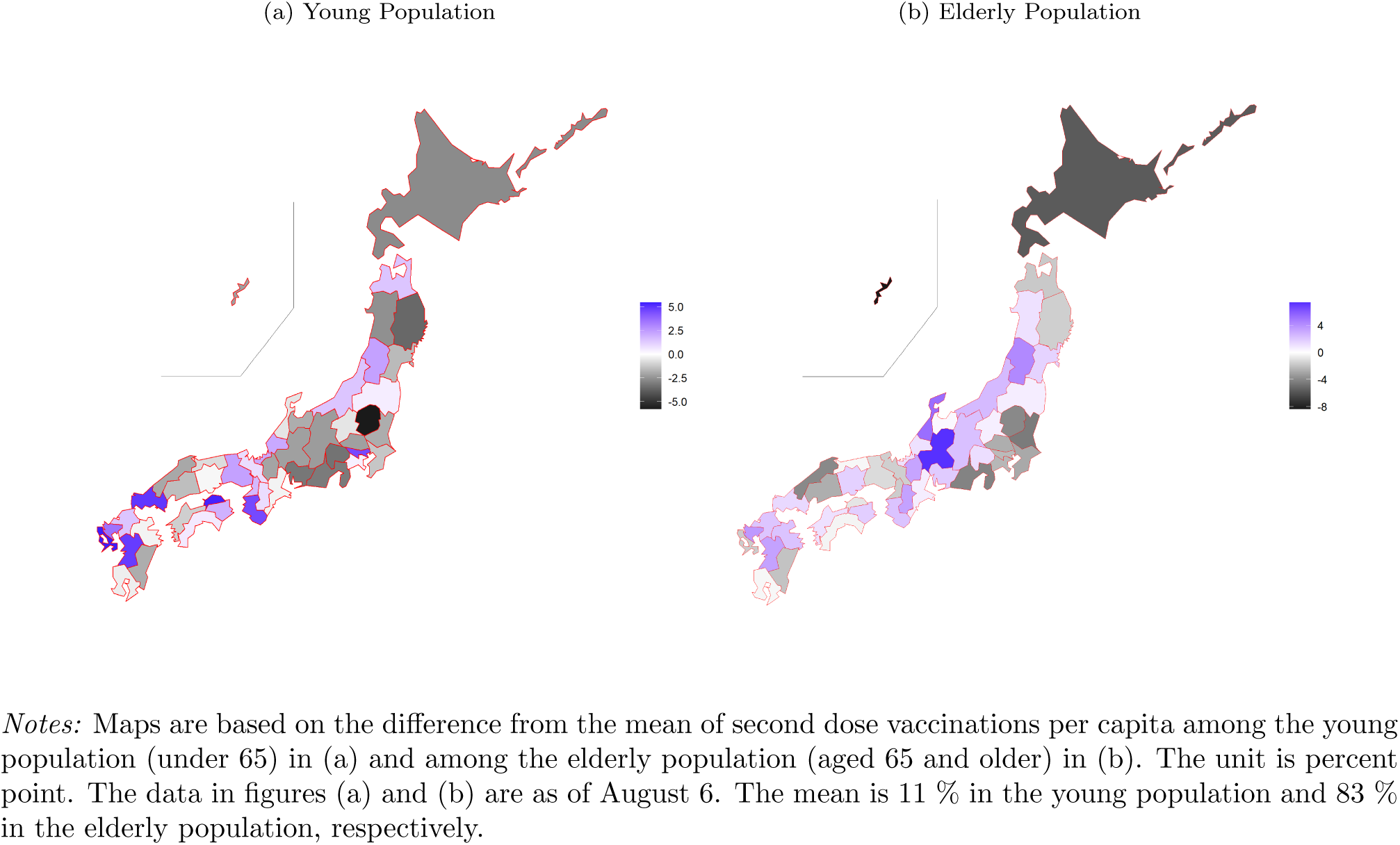
Regional Disparity in Vaccination Rate

**Figure A2:**
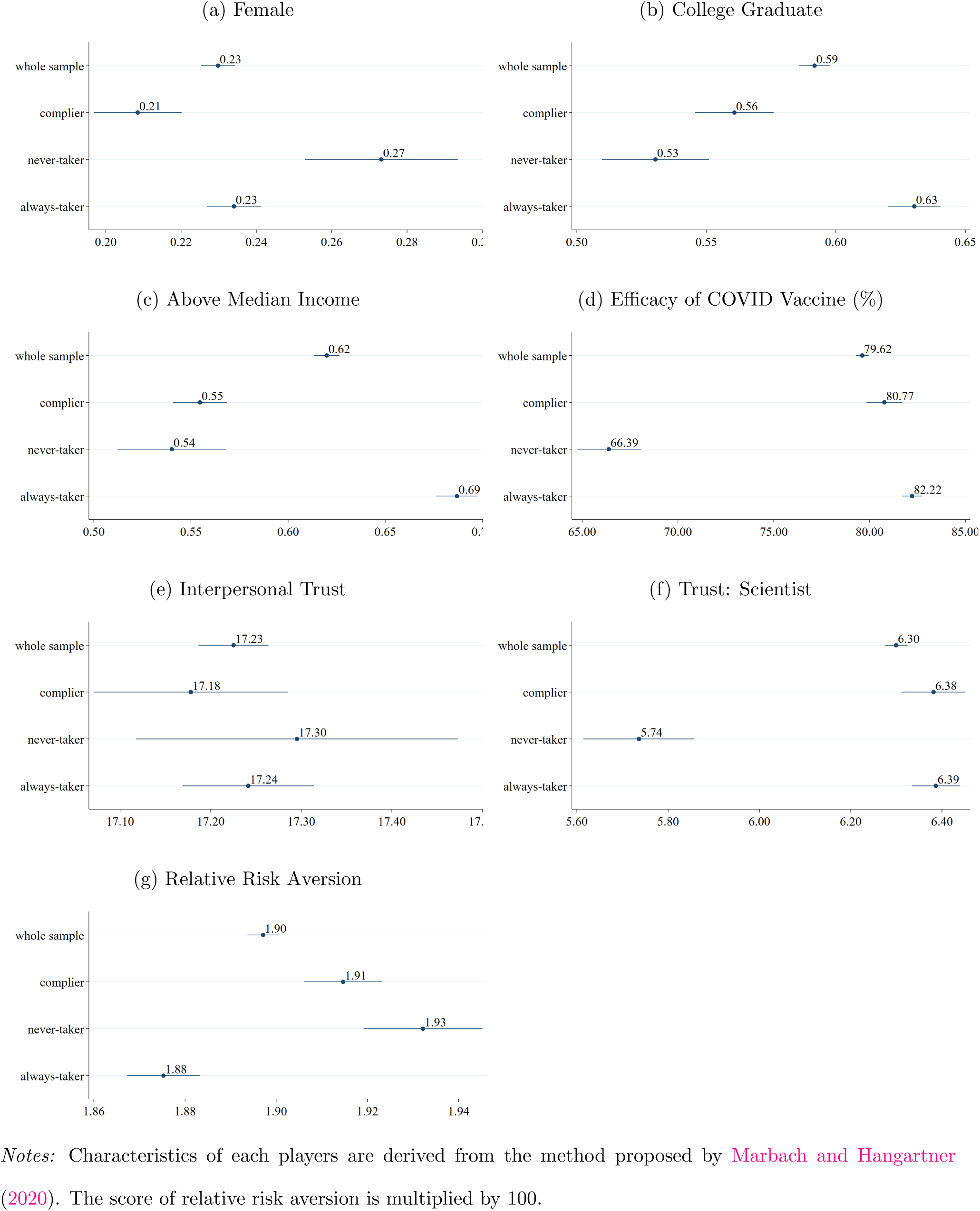
Profile of Each Players

## B Robustness Checks

### B.1 Specification with Covariates

This section presents estimation results controlling for gender, income, employment, education level, presence of chronic diseases, and living in highly infected areas. Appendix Table B1 corresponds to Table 2. The sample size of this estimation is smaller than that of the main results because about 20% of respondents do not provide their income information. Estimates in Appendix Table B1 indicate that our main empirical findings are robust to changes in regression specification. Although estimates for infection control measures by municipal governments are slightly less significant, COVID-19 vaccination increases favorable opinions of the municipal government policies against the pandemic. The estimates also show that COVID-19 vaccination does not affect support for policies by the central government.

Appendix Figure B1 and B2 respectively corresponds to Figure 4 and 5. These estimates show that even when we control covariates, people with chronic diseases and high interpersonal trust are more likely to increase political support due to vaccination. Appendix Figure B3 corresponds to Figure 6 and shows that, as with the main result, the political impact of vaccination is more pronounced among people with low SES. Although the comparison by the regional risk shows ambiguous results, this result reinforces the argument that individual health risk is more important for the political impact of vaccination than regional infection risk. Appendix Table B2 and Appendix Figure B4 respectively correspond to Table 4 and Figure 7. COVID-19 vaccination does not significantly affect people’s health behaviors and mental health, and the difference in estimates by risk preference is small. Overall, our robustness checks replicate the main results that both reciprocal motive and political information can be important factors of the political impact of COVID-19 vaccination.

**Table B1:**
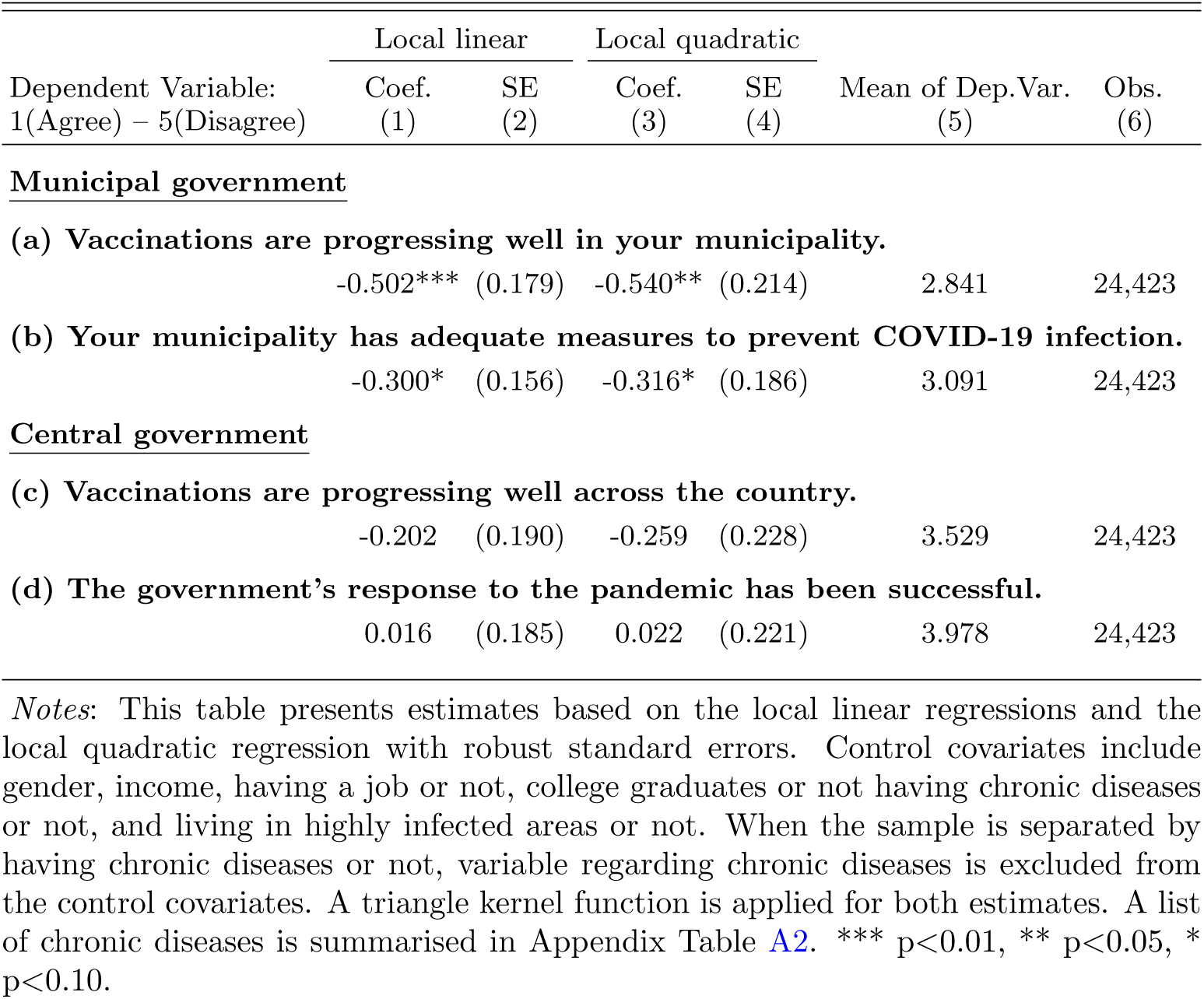
Estimates for the Political Impact of COVID-19 Vaccination (Control Covariates)

|  | Local linear |  | Local quadratic |  |  |  |
| --- | --- | --- | --- | --- | --- | --- |
| Dependent Variable: | Coef. | SE | Coef. | SE | Mean of Dep.Var. | Obs. |
| 1(Agree) – 5(Disagree) | (1) | (2) | (3) | (4) | (5) | (6) |
| <u>Municipal government</u> |  |  |  |  |  |  |
| (a) Vaccinations are progressing well in your municipality. |  |  |  |  |  |  |
|  | -0.502*** | (0.179) | -0.540** | (0.214) | 2.841 | 24,423 |
| (b) Your municipality has adequate measures to prevent COVID-19 infection. |  |  |  |  |  |  |
|  | -0.300* | (0.156) | -0.316* | (0.186) | 3.091 | 24,423 |
| <u>Central government</u> |  |  |  |  |  |  |
| (c) Vaccinations are progressing well across the country. |  |  |  |  |  |  |
|  | -0.202 | (0.190) | -0.259 | (0.228) | 3.529 | 24,423 |
| (d) The government's response to the pandemic has been successful. |  |  |  |  |  |  |
|  | 0.016 | (0.185) | 0.022 | (0.221) | 3.978 | 24,423 |

*Notes:* This table presents estimates based on the local linear regressions and the local quadratic regression with robust standard errors. Control covariates include gender, income, having a job or not, college graduates or not having chronic diseases or not, and living in highly infected areas or not. When the sample is separated by having chronic diseases or not, variable regarding chronic diseases is excluded from the control covariates. A triangle kernel function is applied for both estimates. A list of chronic diseases is summarised in Appendix Table A2. \*\*\* p<0.01, \*\* p<0.05, \* p<0.10.

**Table B2:**
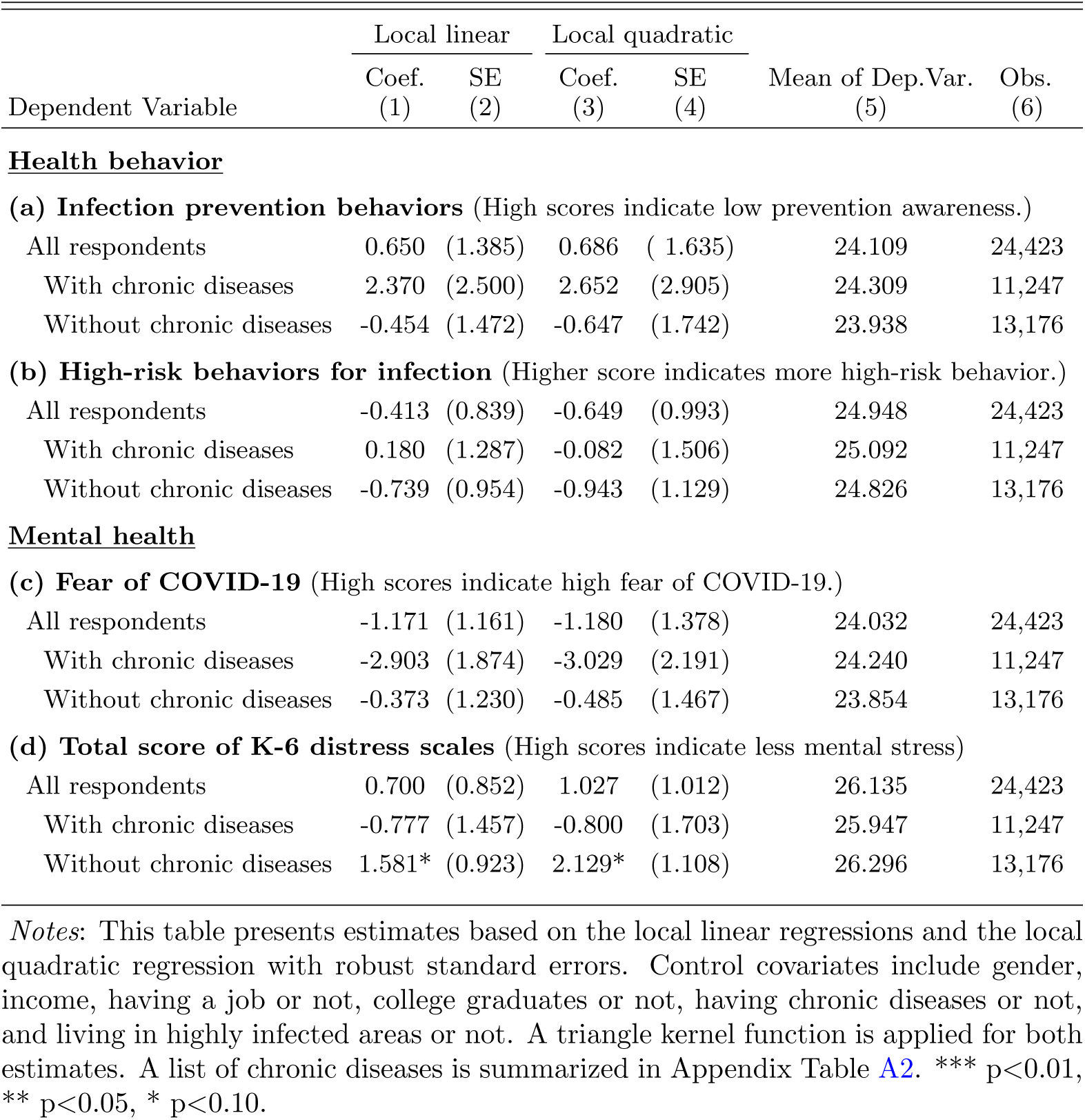
The Impact of COVID-19 Vaccination on Health Behaviors and Mental Health (Control Covariates)

| Dependent Variable | Local linear |  | Local quadratic |  | Mean of Dep.Var.<br>(5) | Obs.<br>(6) |
| --- | --- | --- | --- | --- | --- | --- |
|  | Coef.<br>(1) | SE<br>(2) | Coef.<br>(3) | SE<br>(4) |  |  |
| **<u>Health behavior</u>** |  |  |  |  |  |  |
| **(a) Infection prevention behaviors** (High scores indicate low prevention awareness.) |  |  |  |  |  |  |
| All respondents | 0.650 | (1.385) | 0.686 | ( 1.635) | 24.109 | 24,423 |
| With chronic diseases | 2.370 | (2.500) | 2.652 | (2.905) | 24.309 | 11,247 |
| Without chronic diseases | -0.454 | (1.472) | -0.647 | (1.742) | 23.938 | 13,176 |
| **(b) High-risk behaviors for infection** (Higher score indicates more high-risk behavior.) |  |  |  |  |  |  |
| All respondents | -0.413 | (0.839) | -0.649 | (0.993) | 24.948 | 24,423 |
| With chronic diseases | 0.180 | (1.287) | -0.082 | (1.506) | 25.092 | 11,247 |
| Without chronic diseases | -0.739 | (0.954) | -0.943 | (1.129) | 24.826 | 13,176 |
| **<u>Mental health</u>** |  |  |  |  |  |  |
| **(c) Fear of COVID-19** (High scores indicate high fear of COVID-19.) |  |  |  |  |  |  |
| All respondents | -1.171 | (1.161) | -1.180 | (1.378) | 24.032 | 24,423 |
| With chronic diseases | -2.903 | (1.874) | -3.029 | (2.191) | 24.240 | 11,247 |
| Without chronic diseases | -0.373 | (1.230) | -0.485 | (1.467) | 23.854 | 13,176 |
| **(d) Total score of K-6 distress scales** (High scores indicate less mental stress) |  |  |  |  |  |  |
| All respondents | 0.700 | (0.852) | 1.027 | (1.012) | 26.135 | 24,423 |
| With chronic diseases | -0.777 | (1.457) | -0.800 | (1.703) | 25.947 | 11,247 |
| Without chronic diseases | 1.581* | (0.923) | 2.129* | (1.108) | 26.296 | 13,176 |

*Notes:* This table presents estimates based on the local linear regressions and the local quadratic regression with robust standard errors. Control covariates include gender, income, having a job or not, college graduates or not, having chronic diseases or not, and living in highly infected areas or not. A triangle kernel function is applied for both estimates. A list of chronic diseases is summarized in Appendix Table A2. \*\*\* p<0.01, \*\* p<0.05, \* p<0.10.

**Figure B1:**
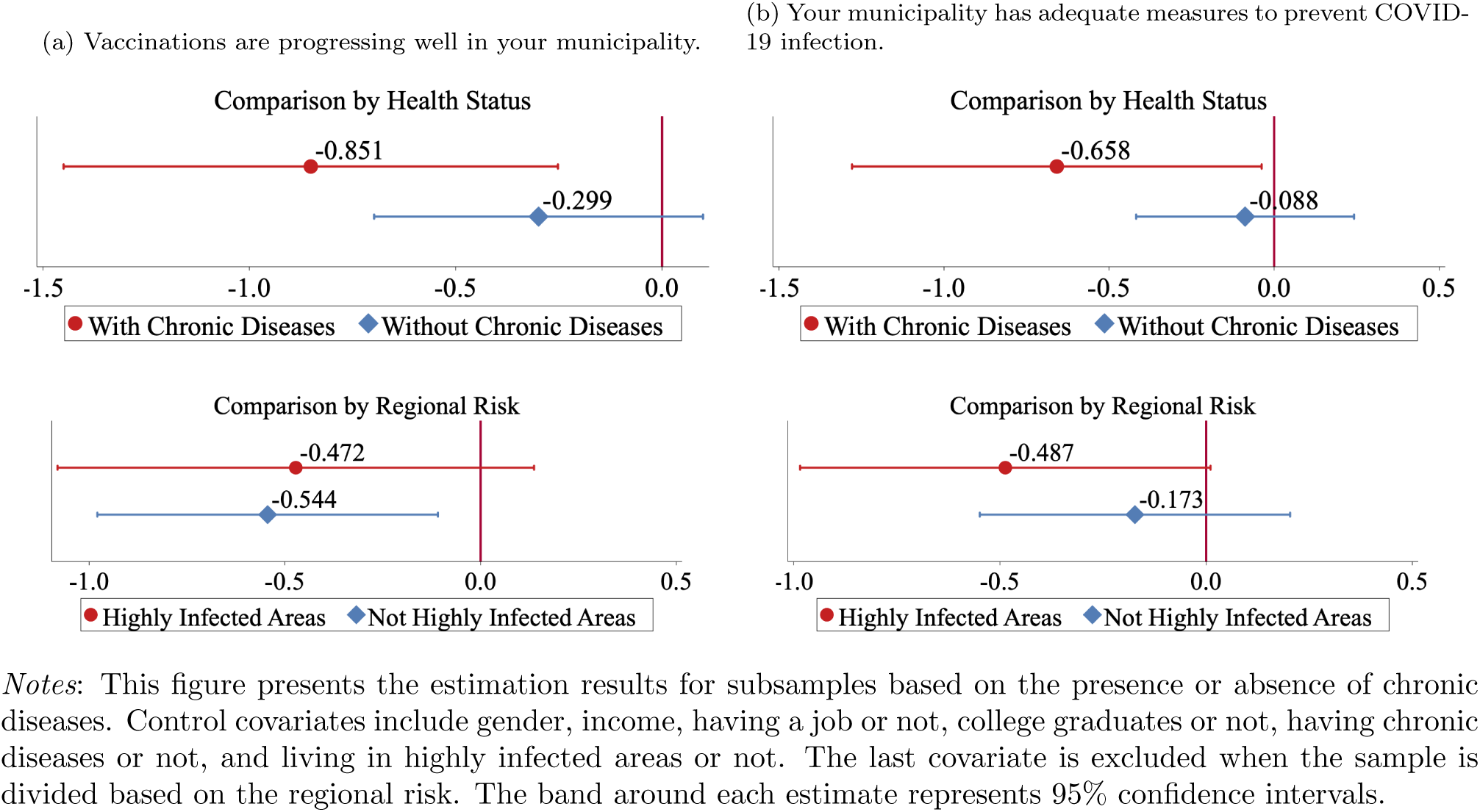
Health Risks and Political Support (Control Covariates)

**Figure B2:**
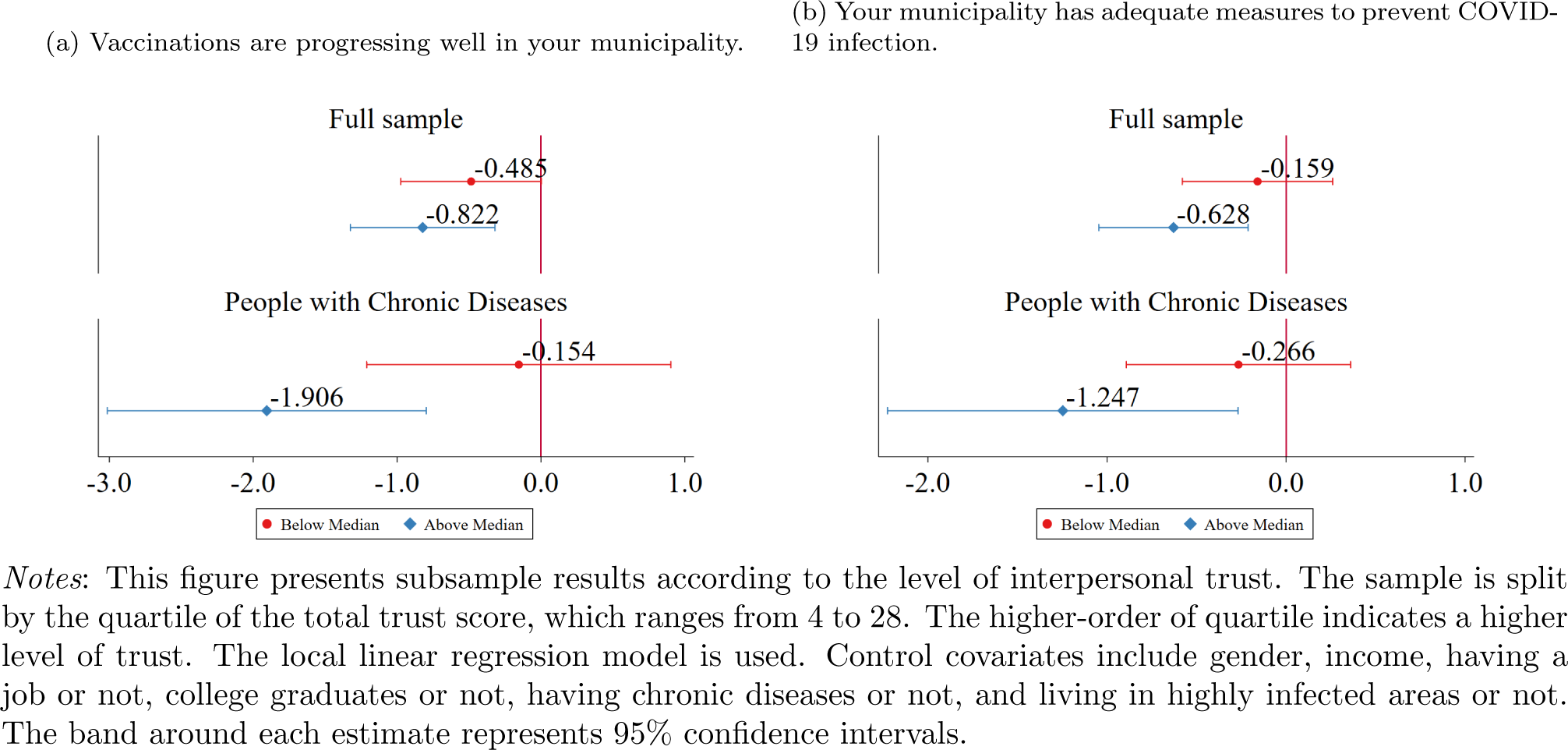
Interpersonal Trust and Political Support (Control Covariates)

**Figure B3:**
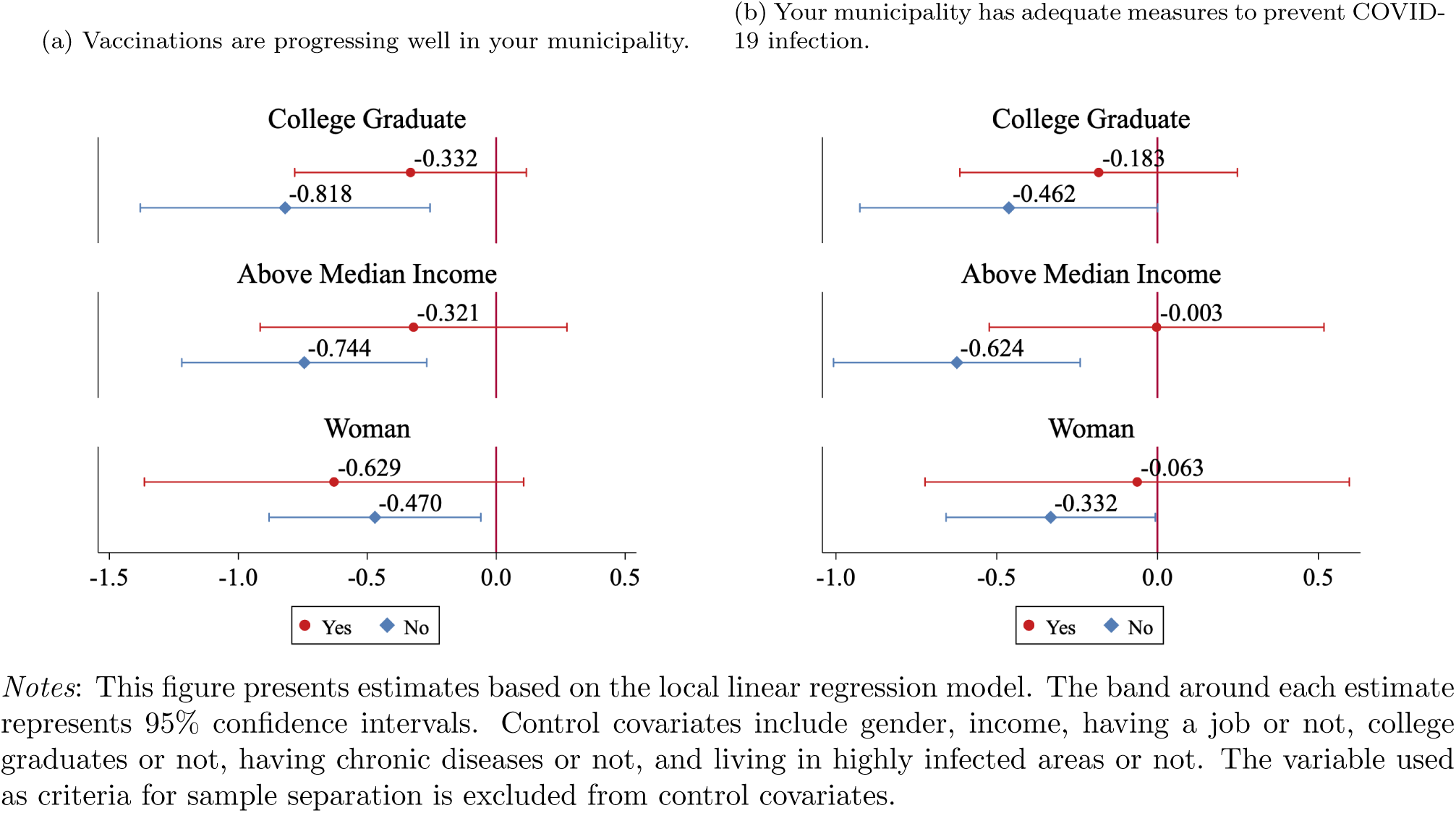
SES and Political Support (Control Covariates)

**Figure B4:**
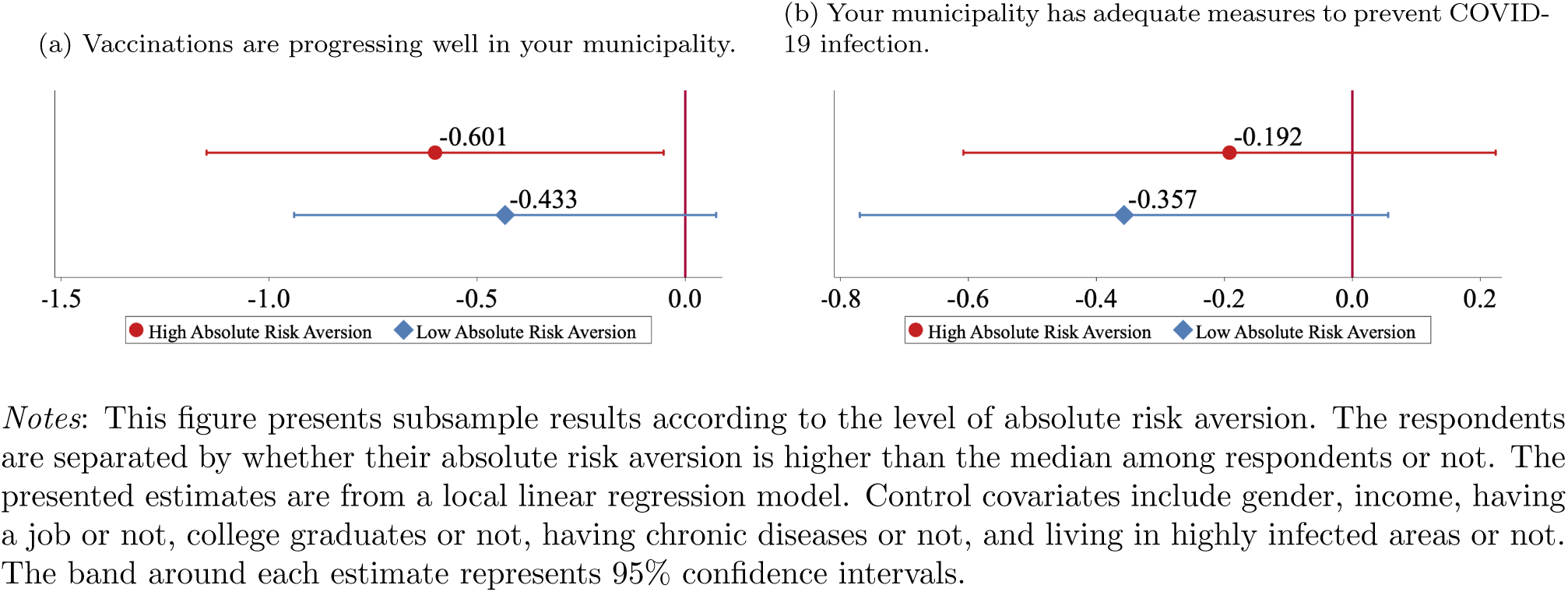
Comparisons by Risk Preferences (Control Covariates)

### B.2 Placebo Test

The results of the placebo tests on vaccination rate and main political outcomes are summarized in Figure B5. Figure Panel (a) in B5 shows the results of vaccination rates. In the horizontal axis, “0” represents the true cut-off month (i.e., April 1, 1957), and the negative(positive) sign means that the threshold is moved to several months before(after) April 1, 1957. Here, we see a large and statistically significant jump when we set the threshold at “0”, indicating that our result on the vaccination rate is not driven by chance.^28^ In addition, we find that RD estimates on the two main outcomes on political support for municipal governments are statistically significant when we adopt the “true” threshold. This also indicates that our results on these outcomes are not explained by the noises and irregular measurement errors around the threshold. We see no statistically significant jumps in all alternative threshold points on the two outcomes on political support for central governments.

Next, we implement the same placebo test for covariates. In continuity tests for covariates in Table A4, we find statistically significant changes in the fraction of college graduates. At the same time, it turns out to be a null effect when we implement Donut-hole RD. Figure Panel (b) in B6 shows that, though RD estimate at “true” threshold is statistically significant, we find many statistically significant results on this covariate. For example, we find statistically significant effects when we set the threshold at “3”, “5”, and “9”. This suggests that data on the fraction of college graduates in this dataset are somewhat noisy, and the statistically significant effects on this variable do not necessarily indicate causal effects.

**Figure B5:**
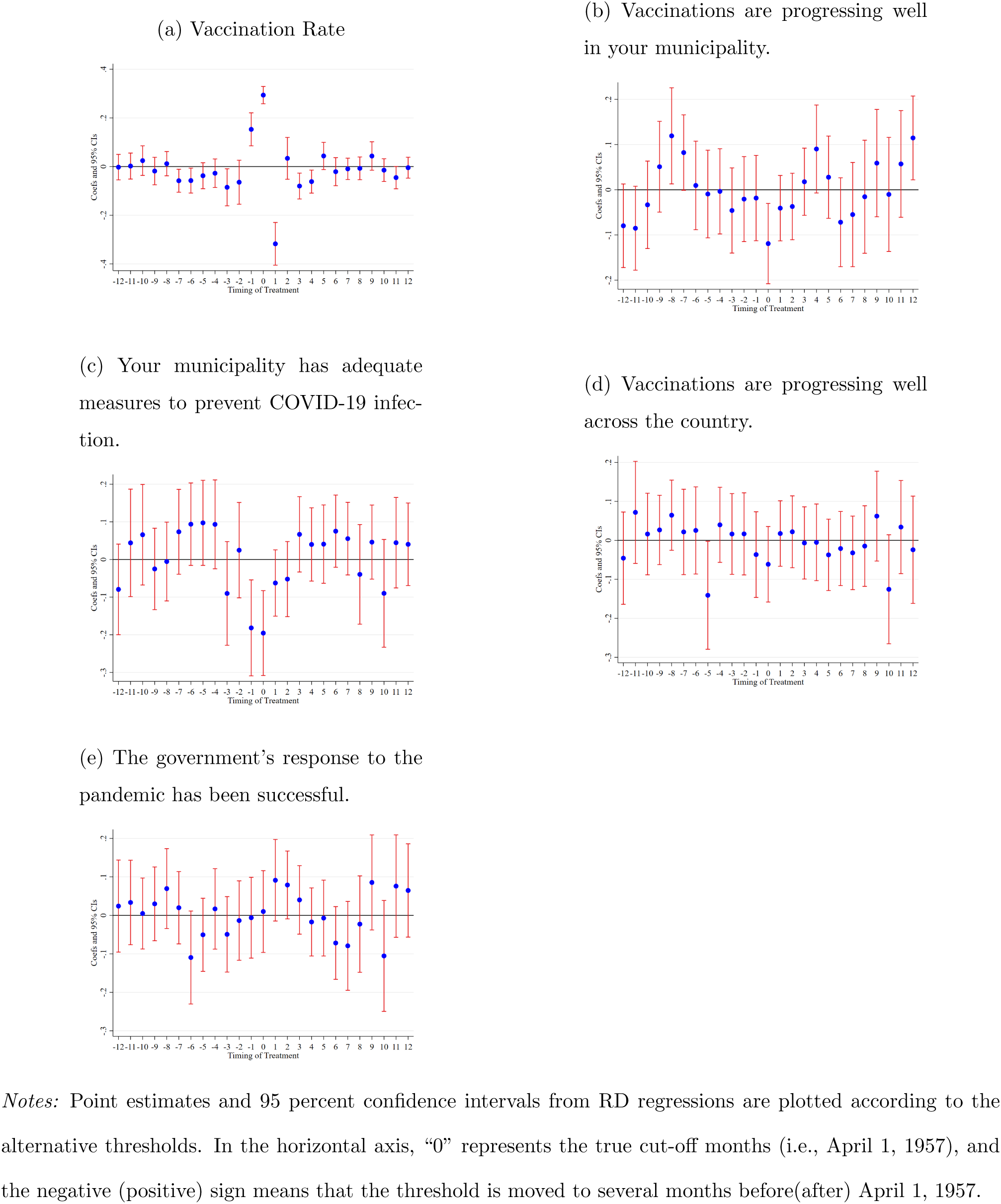
Vaccination Rate and Political Outcomes

**Figure B6:**
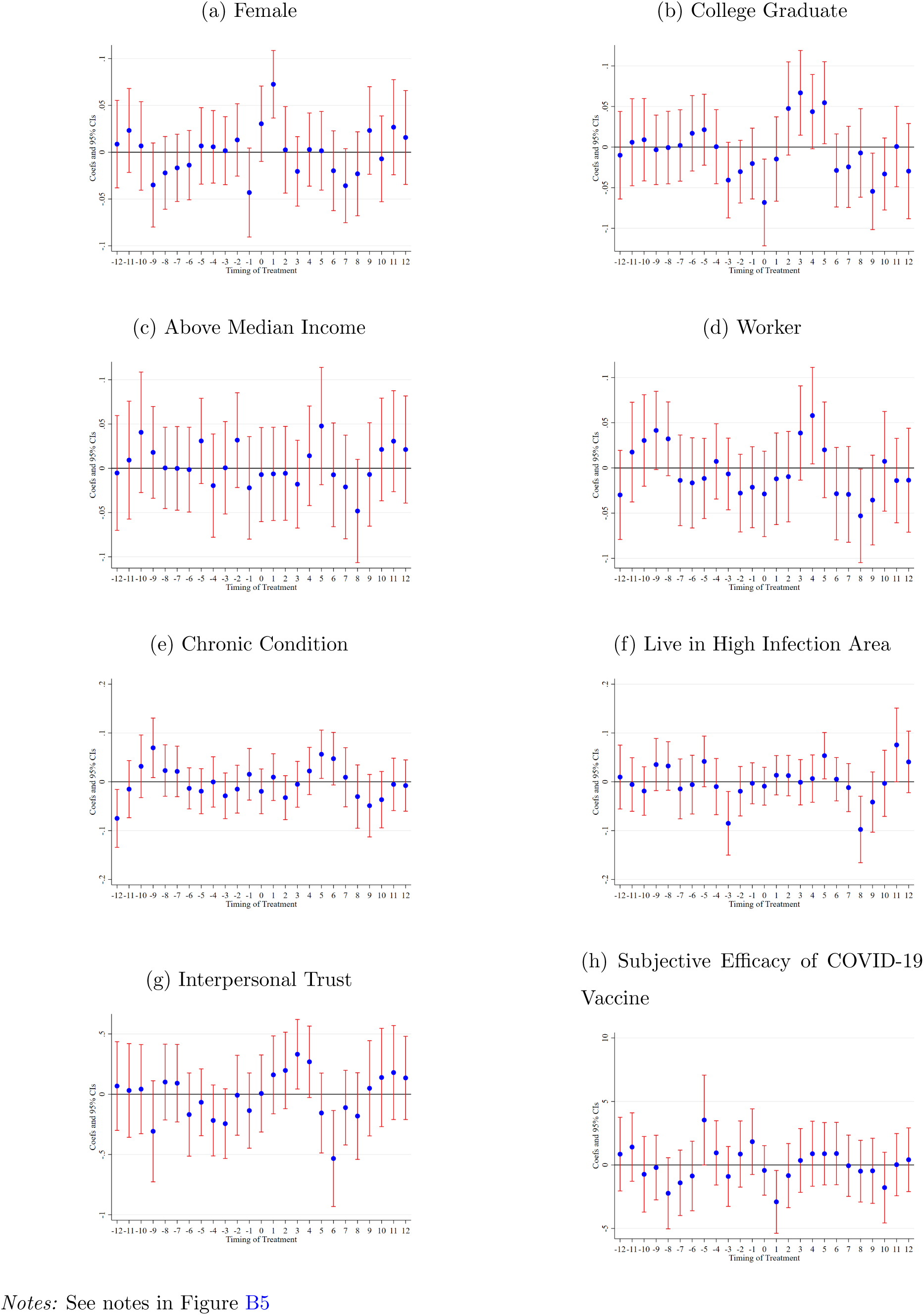
Covariates

## Footnotes

1 The political impact of policies against the pandemic has not been well analyzed, but there are some exceptions. For example, Bol et al. (2021) found that, in Western Europe, lockdown policies had increased support for ruling parties, trust in government, and satisfaction with democracy. Londoño-Vélez and Querubin (2020) also revealed that, in Colombia, cash transfers to poor households during the pandemic increased their support for emergency assistance to households and firms.

2 Trust can be broadly defined as “the expectation that arises within a community of regular, honest, and cooperative behavior, based on commonly shared norms, on the part of other members of the community” (Fukuyama, 1996), which reduces conflict between agents. “Trustworthiness” is related to the reciprocity of a person.

3 Pop-Eleches and Pop-Eleches (2012) analyze the political implications of distributing coupons to poor people to buy computers in Romania.

4 Manacorda et al. (2011) is careful to point out that their results do not rule out behavioral economic explanations.

5 The first mRNA vaccine by Pfizer-BioNTech was approved on Februry 14. In addition, vaccines by Moderna and AstraZeneca were approved on May 21.

6 Although many countries have made vaccines a priority for high-risk populations such as elderly people, priority rules were not strictly enforced. For example, the US government categorized people into three priority groups for vaccination based on age (i.e., 75+ years, 65-74 years, 16-64 years). Still, there is a wide range of regional variation in the actual implementation of this policy (Dooling, 2021). As a result, even in the first month since COVID-19 vaccines were authorized, the share of persons aged less than 65 years who were vaccinated reached 70 percent (Painter et al., 2021), indicating that priority rules were not strictly enforced.

7 In Japan, the fiscal year starts in April and ends in March.

8 To achieve this goal, the central government encourages municipalities to offer high subsidies for health care workers for vaccine shots. While there are no comprehensive statistics, it is well known that the daily salary of physicians for vaccine shots was 1,750 USD in a municipality in rural areas. The daily salary was generally higher in rural areas than in urban areas because of the shortage of human resources.

9 To expand vaccination to the younger generation, the Japanese government also introduced a system of Vaccination in the Workplace (VIW) that allows companies and universities to vaccinate their members.

10 Unlike the lockdowns in other countries, Japan’s regulations under the state of emergency are not legally binding. Instead of mandatory restrictions on behavior, the government requested people to refrain from going out, to limit gatherings, and to shorten the hours of operation of restaurants. On the details of Japan’s state of emergency, see Watanabe and Yabu (2020).

11 The local government system in Japan consists of two tiers: prefectures and the municipalities that make up the prefectures. Municipalities are local public entities that have a close relationship with residents and handle affairs directly related to them. As of 2021, there are 1,718 municipalities in Japan.

12 With survey information on the experience on vote-buying and experimental data on individual intrinsic reciprocity, Finan and Schechter (2012) find politicians can overcome commitment problems under secret ballots by targeting reciprocal individuals.

13 In general, trust is understood as a more general concept than reciprocity because why people trust someone else can also be explained by pure altruism (Cox, 2004) as well as reciprocal motivation. The distinction between trust through generosity and trust through reciprocity is not so important in our study because, from either motivation, voters are likely to be grateful for the government when they receive public services.

14 Son^g^ (2008) also conducted a laboratory experiment and show that trusting behavior is driven by reciprocity expectations. Dohmen et al. (2008) explore the determinants of trust and reciprocity using individual survey data and find a strong correlation between them.

15 A limitation of our analysis is that due to the policies of the survey company, we were unable to ask respondents whether they support the central government or the municipal government itself. Therefore, we could not ask what respondents thought of a description such as “You support the Japanese government (or the municipal government in which you live).” We instead measure support for the government by asking opinions about their policies against the pandemic.

16 The original version in Siegrist and Bearth (2021) uses two other questions about trust in strangers are added. However, we did not use these two questions because, in our context, trust in the people in the same locale rather than strangers is important.

17 This is because, only in the third questionnaire item, lower values indicate higher interpersonal trust.

18 The trust in scientists is measured by a 10 point scale as is in the European social survey (Helliwell et al., 2016) (0 ”Not Trust at all” - 10 ”Completely Trust”).

19 Note that the subjective vaccine efficacy of the respondents is much lower than the official data from Pfizer and Moderna (about 95%).

20 Even when the mandatory retirement age is 65 years in some companies, the timing of retirement is the end of the fiscal year in which they turn 65. Therefore, the age 65 threshold of vaccination is one year earlier than the timing of mandatory retirement.

21 In Online Appendix B.2, we also implement placebo tests by changing the threshold point. As for the fraction of college graduates, RD estimates from some fake thresholds are statistically significant. This also suggests that data on college graduates are somewhat noisy and include irregular values.

22 In the actual implementation, we use the STATA package developed by Marbach and Hangartner (2020).

23 The score of compliers and never-takers is 6.38 and 5.72, respectively. Nomura et al. (2021) also find that trust in scientists is associated with the unwillingness to take the COVID-19 vaccine.

24 As highly infected areas, we choose the following prefectures: Tokyo, Kanagawa, Saitama, Chiba, Aichi, Kyoto, Osaka, and Fukuoka.

25 Infection prevention behaviors are the total score of responses to questionnaires in “Daily Preventive Behavior,” and high-risk behaviors for infection are the total score of responses to questionnaires in “Going-out Behavior” in Appendix Table A1.

26 The questionnaire items on risk preferences are the same as those used in the Japan Household Panel Survey on Consumer Preferences and Satisfaction (JHPS-CPS). See Hanaoka et al. (2018) for details.

27 The Arrow-Prat measure of absolute risk aversion is calculated as 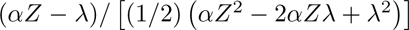, where we denote *α* as the probability of willing lottery, *Z* as the prize of lottery, *γ* as the willingness-to-pay amount.

28 When we set the threshold at “1”, we find large and negative effects. This is because the trend of vaccination rate becomes very steep at the left-hand side of this fake threshold.

